# Reproducible ecological restructuring of the oral microbiome defines the oral-gut axis in Crohn’s disease

**DOI:** 10.64898/2026.04.28.26351936

**Authors:** RJ. Whelan, DIF. Wands, P. Rimmer, R. Hansen, DC. Wilson, G. Moran, J. Imai, GL. Hold, ILC. Chapple, T. Iqbal, K. Gerasimidis, GT. Ho

## Abstract

The oral microbiome is increasingly implicated in inflammatory bowel disease (IBD), yet findings across studies remain inconsistent and difficult to reconcile due to methodological heterogeneity. To resolve this, we performed a cross-cohort integration of oral microbiome datasets across 25 studies (n = 1,136 IBD; n = 759 controls), combined with unified re-analysis of publicly available sequencing data and newly generated in-house data within a single bioinformatic framework. Oral microbial diversity was consistently reduced in IBD (standardised mean difference –0.31, p = 0.007), with replication in the harmonised dataset (Hedges’ SMD = –0.372, p < 0.001), driven predominantly by Crohn’s disease (CD). Multivariable modelling identified a reproducible set of genera associated with disease, including enrichment of *Corynebacterium*, *Serratia* and *Streptococcus*. Network analysis demonstrated a fundamental ecological shift: whereas healthy communities were organised around a *Selenomonas* hub, CD exhibited integration of *Corynebacterium* into core network architecture, consistent with a shift toward an inflammation-adapted, aerotolerant ecological state. Functional predictions supported this restructuring, with depletion of butyrate metabolism and enrichment of aromatic amino acid degradation pathways. Together, these findings identify reproducible ecological restructuring of the oral microbiome in CD and establish a cross-cohort framework linking oral microbial organisation to the oral-gut axis in IBD.

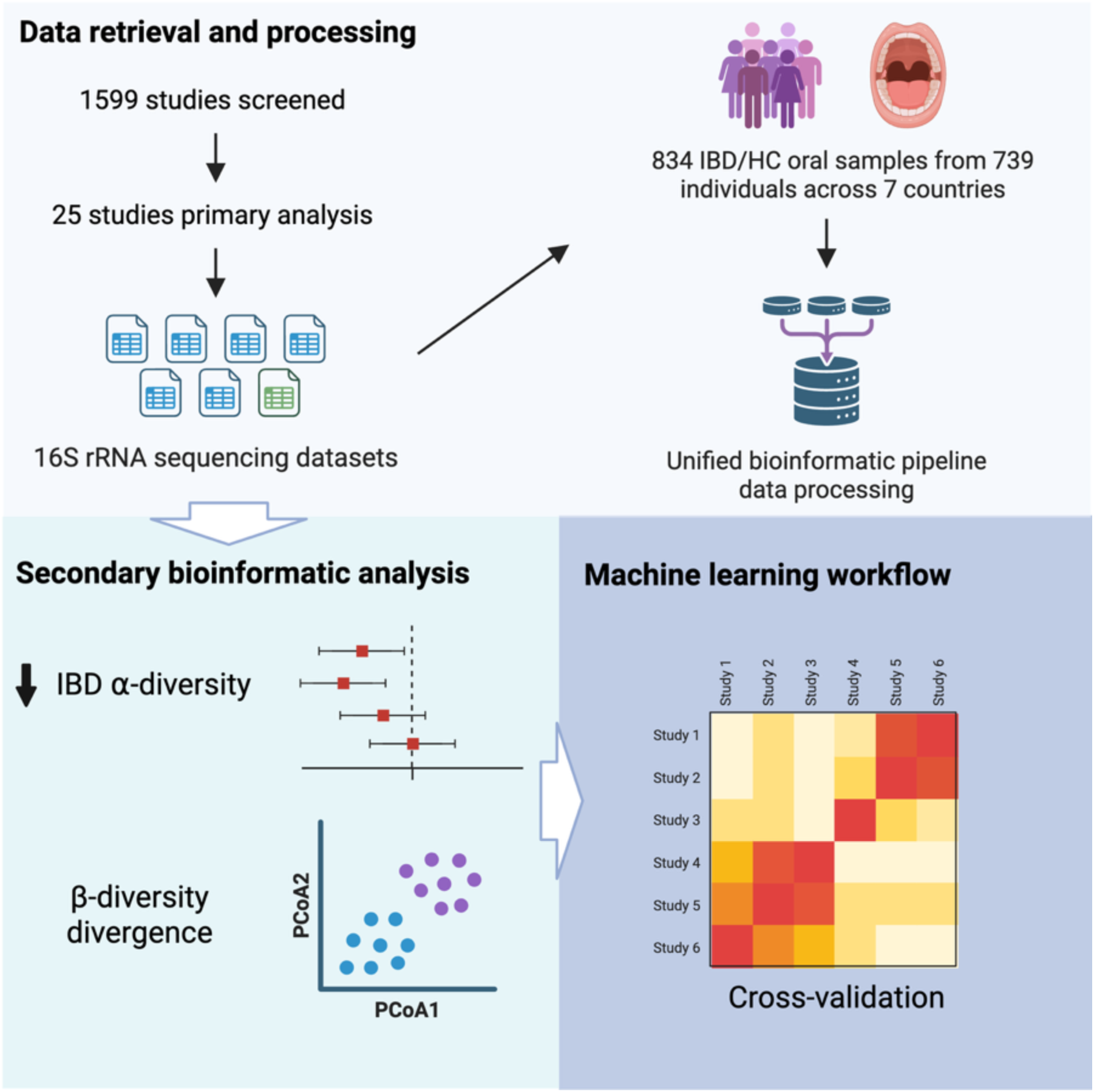

## Introduction

Inflammatory bowel disease (IBD) is a chronic, immune-mediated disorder of rising global incidence^1,2^ in which the gut microbiome is now recognised as a major factor in disease onset and progression.^3–5^ Microbiome-based approaches are being increasingly explored and harnessed for diagnosis, risk prediction, and therapeutic intervention.^6,7^ A growing and convergent body of experimental, ecological and clinical evidence argues that the upstream compartment, the mouth, is not a passive reservoir but an active, mechanistically relevant component of the de-regulated host-bacteria interaction in IBD.^8,9^

The oral cavity is the second most complex microbial ecosystem in the human body and the anatomical gateway to the gut. A healthy adult produces and swallows approximately 1.5 litres of saliva each day, delivering a continuous stream of oral microbes to the stomach, small intestine and colon.^10,11^ The oral microbiome is the conduit for initial gut microbial colonisation at birth, its composition differing between vaginal and Caesarean deliveries;^12^ and continues to seed and influence distal gut communities throughout life.^13^ Diet, alcohol (particularly refined sugars) and smoking, all established IBD environmental factors directly modulate oral microbial composition.^14,15^

The oral mucosa is intrinsically primed for inflammation. A recent single-cell atlas of human oral mucosa demonstrated that stromal and immune populations are transcriptionally programmed to recruit neutrophils, embedding inflammatory potential within resident tissue biology.^16^ Experimental work has provided causal evidence that oral bacteria can directly drive Crohn’s disease (CD)–relevant immunity: transplantation of saliva from patients with CD into germ-free mice induces IFN-γ⁺ Th1 accumulation in the intestinal lamina propria, with the oral-origin pathobiont *Klebsiella pneumoniae* 2H7 recapitulating colitis in genetically susceptible *Il10*^−/−^ hosts.^17,18^ Oralisation, the ectopic colonisation of the gut by oral bacteria is supported by several studies that show enrichment of oral taxa within inflamed intestinal mucosa in IBD^3,19,20^ and by strain-level concordance between oral and tumour-associated *Fusobacterium nucleatum* in colorectal cancer.^21^

Inflammation also creates a permissive niche for ectopic oral pathobionts in the gut, establishing a feed-forward oral–gut loop.^22^ Periodontal inflammation simultaneously generates gut-tropic, oral pathobiont-reactive Th17 cells that migrate to the inflamed intestine, where they are activated by translocated oral bacteria and drive colitis.^18^ Oral inflammation therefore exacerbates gut disease by supplying both colitogenic microbes and pathogenic immune effector cells. Parallel work places *Fusobacterium nucleatum*, *Campylobacter concisus* and periodontitis-derived pathobionts in the gut Th17 immune response.^21,23–26^

The oral compartment is accessible and non-invasive for mucosal microbial sampling,^8^ yet the human oral microbiome literature in IBD remains fragmented: single-centre cohorts, heterogeneous sampling sites (saliva, tongue, buccal, plaque), inconsistent bioinformatic pipelines, and non-overlapping lists of differentially abundant taxa. Here we address this gap by performing the largest harmonised analysis of the oral microbiome in IBD to date, integrating primary 16S rRNA sequencing datasets across multiple independent cohorts within a unified bioinformatics framework. Rather than focusing solely on differential taxonomic abundance, we interrogate whether reproducible ecological structures emerge across studies despite methodological heterogeneity. Specifically, we sought to (i) define cross-cohort alterations in oral microbial diversity and composition across IBD subtypes; (ii) identify taxa and ecological interactions that persist following multivariable adjustment; (iii) determine whether community-level network architecture is conserved or restructured in disease; and (iv) infer functional adaptations associated with these ecological states. Through this approach, we aim to establish a reproducible, systems-level framework for understanding the oral microbiome in IBD and its role in the oral-gut axis.

## Materials and methods

### Scotland IBD oral microbiome cohort

The MUSIC study is a prospective, multicentre, longitudinal cohort conducted in real-world IBD clinical care (www.musicstudy.uk). The study was approved by the East of Scotland Research Ethics Service (REC 19/ES/0087), and all participants gave written informed consent. Oral salivary DNA were collected as part of the biomarker profiling approach to predict mucosal healing in IBD.

Participants were asked to complete an oral health questionnaire at sampling visit to record any confounding variables, including mouth piercings, frequency of tooth brushing and presence of any oral manifestations. Participants were asked to refrain from tooth brushing, eating, drinking and smoking one hour prior to saliva sampling. An unstimulated active saliva spit was collected in an OMNIgene ORAL 505 tube DNA (Genotek Incorporation, Ontario, Canada) containing a rapid microbial DNA and RNA stabilisation solution. Samples were subsequently processed following the manufacturer’s instructions and stored at-80°C.

### Microbial DNA extraction and 16S rRNA sequencing of the oral Scotland IBD cohort

Microbial DNA from saliva samples was extracted using the DNeasy Powersoil Pro kit (Qiagen) following the manufacturer’s instructions and using the TissueLyser II for 10 minutes at 25 Hz for mechanical bead-beating and cell lysis. As per the manufacturer’s instructions, the starting sample volumes used for saliva was 250 µl. A negative and positive (Zymobiomics Microbial Mock community standard) control sample were included in each extraction batch.

The 16S rRNA sequencing libraries were prepared and sequenced by NU-OMICS DNA Sequencing facility (Northumbria, UK) using the Schloss lab protocol.^27^ Briefly, the V4 region of the 16S rRNA gene was firstly amplified using the following primers (515F: GTGCCAGCMGCCGCGGTAA and (806R (GGACTACHVGGGTWTCTAAT)) with Illumina overhang adapter sequences.^28^ A second round of PCR attached Illumina dual-index barcodes for sample multiplexing. Sequencing was performed on the MiSeq system (Illumina) producing 250 bp paired-end reads. A negative and positive (Zymobiomics Microbial Mock community DNA standard) control sample were included in each 96-well plate and carried through to sequencing.

### Protocol and registration

This systematic review follows the criteria outlined in the Preferred Reporting Items for Systematic Reviews and Meta-analyses (PRISMA) statement.^29^ The protocol is registered on PROSPERO (#CRD42024573554) and can be accessed at: https://www.crd.york.ac.uk/prospero/display_record.phptiID=CRD42024573554.

### Eligibility criteria

Study selection was restricted to only peer-reviewed, full-text studies. Study inclusion criteria were: (1) observational study designs including cross-sectional, case-control or intervention studies, (2) all patients with a diagnosis of IBD based on clinical, endoscopic, histological and radiological findings, including adults and children, (3) reporting of oral microbiota diversity and composition data using 16S ribosomal RNA and/or shotgun metagenomic sequencing. Exclusion criteria were (1) non-use of next-generation sequencing technology, (2) studies that did not present IBD alongside a comparator group of controls (either healthy or symptomatic non-IBD).

### Information sources and search strategy

Study search was performed using the following platforms: Embase via OVID, Web of Science (limited to science citation index), Medline via OVID Cochrane and SCOPUS. The search included articles published from database inception to 1^st^ April 2025 using the search outlined in the PROSPERO protocol (#CRD42024573554).

### Selection and data collection process

Covidence^30^ was used for duplicate removal, study screening, and abstract review. Full-text screening was performed independently by two reviewers (RW and DW). Conflicts in study inclusion decisions were resolved through discussion until a consensus was reached with GTH as the moderator. Two reviewers (RW and DW) extracted the data from eligible studies.

### Data items

The extracted dataset included publication information, participant demographics, disease phenotype, type of control, sampling method and criteria, microbiome sequencing and analysis methods, microbiome results including diversity metrics, relative abundance analysis and functional prediction, and sequencing and metadata availability and location. Any missing data is noted in the summary tables (S1 and S2).

### Study risk of bias assessment

The Newcastle-Ottawa quality assessment scale for case-control studies was used to assess the internal validity and risk of bias of included studies.^31^ A quality score maximum of 9 stars was assigned to each study based on the following domains: study selection, the comparability of the groups and the ascertainment of the exposure. Of note, the main confounder variables highlighted were the inclusion of a dental exam, the use of antibiotics one month before sampling and patient fasting before sampling.

### Data retrieval

Shannon diversity metrics were extracted from papers either as presented numerical values or inferred from graphs. Values were inferred from graphs using WebPlotDigitizer version 4.7.^32^ Studies chosen for secondary analysis had to use 16S rRNA sequencing technology and be performed on an Illumina platform. Shotgun metagenomic sequencing data and data sequencing via pyrosequencing were excluded from the analysis as they were not compatible for our unified analytical approach. Raw sequencing data from studies were downloaded via the fetchngs nf-core pipeline version 1.12.0 using the respective study identifier number, SRA or otherwise.^33^ Where data was not available publicly or datasets contained missing data, study authors were contacted for data availability. We only included patient baseline samples for studies with longitudinal sampling. Periodontist labelled samples from PRJNA684508 were removed from downstream analysis. The control group for all studies were from a healthy population except SRP385133 which were symptomatic controls.

### Data processing and ASV assignment

Fastq files were manually inspected for the presence of primers using the R package Biostrings (v2.72.1). Cutadapt (v5.0) was used to remove primers using in-house scripts. Data were processed using nf-core ampliseq version 2.14.0 of the nf-core collection of workflows, utilising reproducible software environments from the Biocontainers projects.^33^ Data quality was evaluated with FastQC^34^ and summarised with MultiQC.^35^ Sequences were processed sample-wise (independent) with DADA2^36^ to eliminate PhiX contamination, trim reads before median quality drops below 30 and ensure at least 80% of reads are retained. Forward reads and reverse reads were trimmed at 250 bp and 225 bp, respectively, and reads shorter than this were discarded. Samples containing less than 10,000 clean reads were removed from downstream analysis. Taxonomic assignment was performed using DADA2 and the expanded human oral microbiome database (16S rRNA RefSeq: V16.02).^37^ Low abundant ASVs were filtered out (<0.01%) of the dataset and the ASV abundance table was normalised using the total sum scaling (TSS) method to relative abundances. Abundances were further collapsed to the genus level for downstream analysis.

### Microbiome data analysis

Shannon diversity standardised mean difference for individual studies was compared between IBD patients (and IBD subtypes) to healthy controls and between high and low IBD disease activity using random effects meta-analysis using RevMan, Version 9.10.0 (The Cochrane Collaboration). For the secondary analysis, study metadata was merged with the phyloseq object produced by the ampliseq pipeline. The phyloseq package (v1.38.0) was used for downstream analysis.^38^ Alpha-diversity analyses were generated using phyloseq and the Shannon and observed diversity indexes were used. Analyses were performed on non-rarefied genus-level data. Beta-diversity analyses were performed with Vegan (v2.6-8) using Bray-Curtis dissimilarity matrices from TSS normalised data. PERMANOVA analysis was performed with Vegan. PICRUSt2 was used to predict altered MetaCyc pathways based on 16S rRNA genus-level data.^39^

We applied the SPIEC-EASI framework to infer microbial association networks.^40^ This accounts for the compositional nature of microbiome data and reduces spurious correlations by estimating sparse, conditionally dependent relationships between taxa. SPIEC-EASI was run using the ‘mb’ method and networks were based on genera that were >5% sample prevalence and >0.01% abundance. Analysis was performed on centred-log ratio transformed matrices. MaAsLin2 multivariate modelling was performed to determine differentially abundant genera and MetaCyc pathways between groups.^41^ Specifically, diagnosis subtype (HC as the reference), sample type, and adult vs paediatric were used as ‘fixed-effects’. ‘Random-effects’ consisted of sample geography (continent) and hypervariable region sequenced. P-values were corrected using the Benjamini-Hochberg method and the threshold for significantly associated taxa was q < 0.05.

### Meta-analysis of alpha diversity

Due to the batch effects that are inherent when analysing data from multiple cohorts, we used a meta-analytical approach using Hedges’ standardised mean difference (SMD).^26^ Hedges’ SMD applies a bias correction to Cohen’s *d* estimator to studies with low sample sizes.

The metacont R package was used to carry out the meta-analysis. Restricted maximum likelihood and confidence intervals (CIs) of the summary effect were used to determine between-study variance and adjusted using the Hartung and Knapp method. Hedges’ SMD was applied to Shannon diversity and richness metrics for studies which had the specific comparisons.

### Machine learning pipeline

We evaluated the performance of random forest classifiers using 1,000 trees, minimum samples per leaf = 2 with hyperparameter tuning of the max_features parameter (sqrt, log2, or unrestricted). Classifiers were trained on TSS normalised oral microbiota genus features, with ASVs >0.01% used. The machine learning environment used the following package versions: scikit-learn (v1.5.2), scipy (v1.14.1), pandas (v2.2.3) and numpy (v2.0.2). Hyperparameters were selected by 3-fold stratified cross-validation on the training set only, and the best model was subsequently evaluated on the held-out test set. Model performance was quantified as the area under the receiver operating characteristic curve (AUC).

We evaluated the random forest classifiers using three different strategies. (1) Within study cross validation: 3-fold stratified cross-validation was repeated 20 times with training and testing performed on the same study dataset. The mean AUC was determined and reported for each study. (2) Across study validation: to test model transferability across studies, classifiers were trained on one study and tested on each other study to produce a study-study performance matrix. (3) Leave-one-dataset-out (LODO): to test model generalisability, we trained models on pooled datasets and tested on held-out dataset and subsequently repeated this for each study. Lastly, overall averages were taken for each of these cross-validation methods.

### Quantification and Statistical Analysis

Statistical tests and data visualisation were performed using R version 4.4.1. Student t-test was performed for two independent groups that were normally distributed. One-way ANOVA followed by Tukey’s HSD post-hoc test for multiple comparisons.

## Data and code availability

Code used to process and analyse the data is available at https://github.com/1-gut/ oral_microbiome_IBD_meta. Bioproject accessions for studies used in the analysis are listed in the resources table.

## Results

### Study selection and harmonised dataset characteristics

Our database search identified 3,111 studies (Embase - 1126, Medline - 695, Web of Science - 613, Scopus - 571, CENTRAL - 106), with 1,599 remaining after the removal of duplicates. Following primary screening, 32 studies were selected for full-text review, of which 25 met the inclusion criteria and were included in the review (Fig. S1). Individual study patient demographics, sample numbers, confounding variables, and microbiome sample collection, storage and analysis are outlined in tables S1 and S2. Of the 25 studies that met the inclusion criteria, 9 did not provide information on data location or availability (Fig. S2). The 6 available 16S rRNA sequencing datasets in addition to our new in-house data included 57 IBDU, 253 CD, 157 UC, and 367 HC samples (Fig. 1B, Table 1). Oral sampling sites varied, comprising 483 saliva, 262 tongue, and 89 buccal. The hypervariable regions targeted were V1–V2 (351 samples), V4–V5 (282 samples), V3–V4 (141 samples), and V4 (60 samples). The analysis included data from six adult studies (423 samples), our in-house dataset (60 samples), and one paediatric study (351 samples).

**Figure 1.**
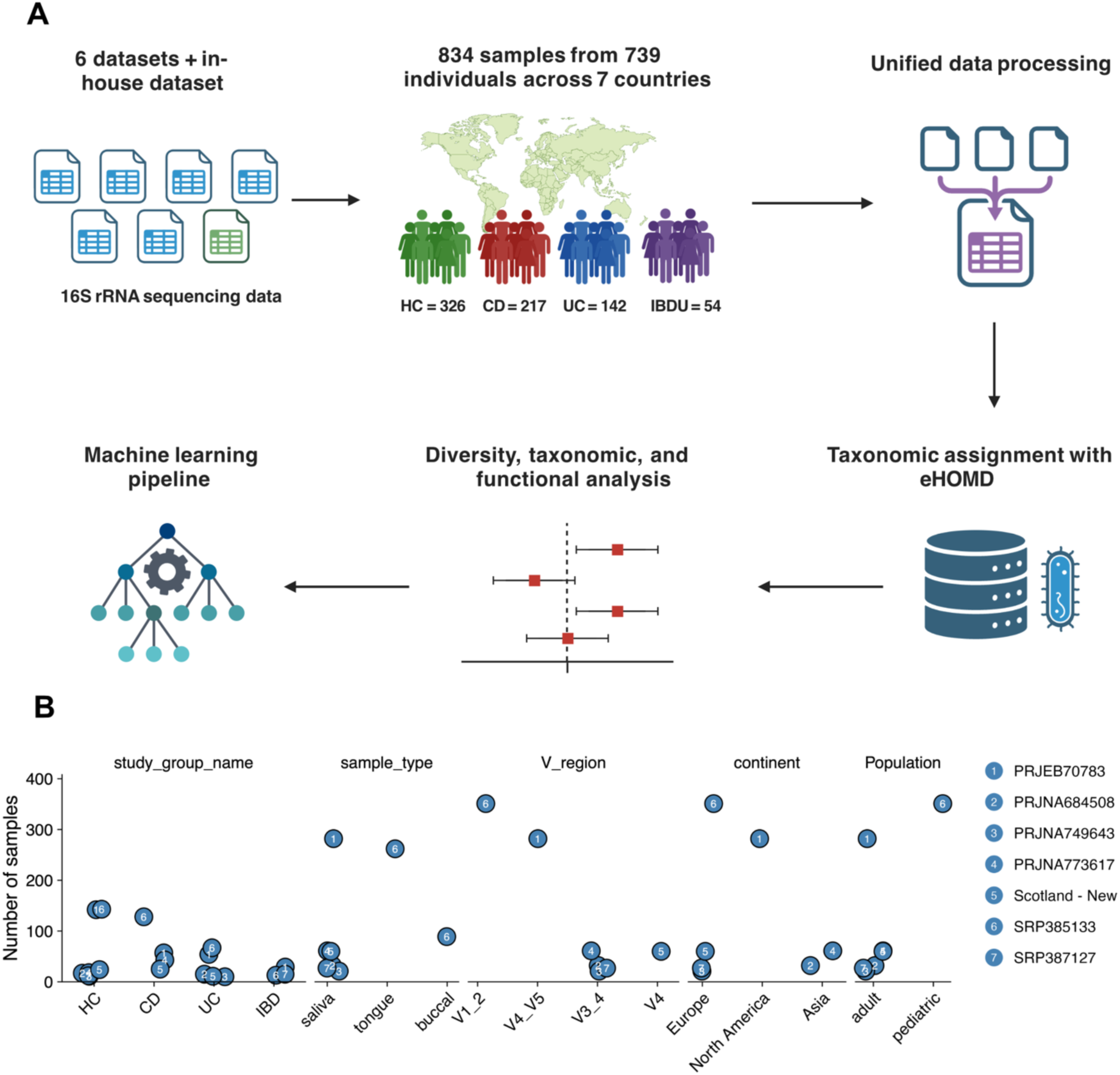
Meta-analysis workflow and study characteristics (A) Workflow of the secondary analysis of study selection, data integration and analysis strategy. Six datasets and our own in-house datasets representing 217 CD, 142 UC, 54 IBDU and 326 HC patients were included in the downstream analysis. Microbial datasets were processed and taxonomy assigned using a unified bioinformatic pipeline and the expanded human oral microbiota database (eHOMD). A meta-analysis was performed to determine community diversity and taxonomic difference between disease groups. Random Forest models were trained to determine the classification potential for IBD and IBD-subtypes. (B) Study sample details across multiple variables including disease phenotype, sample type, hypervariable region sequenced, continent and study population.

**Table 1.**
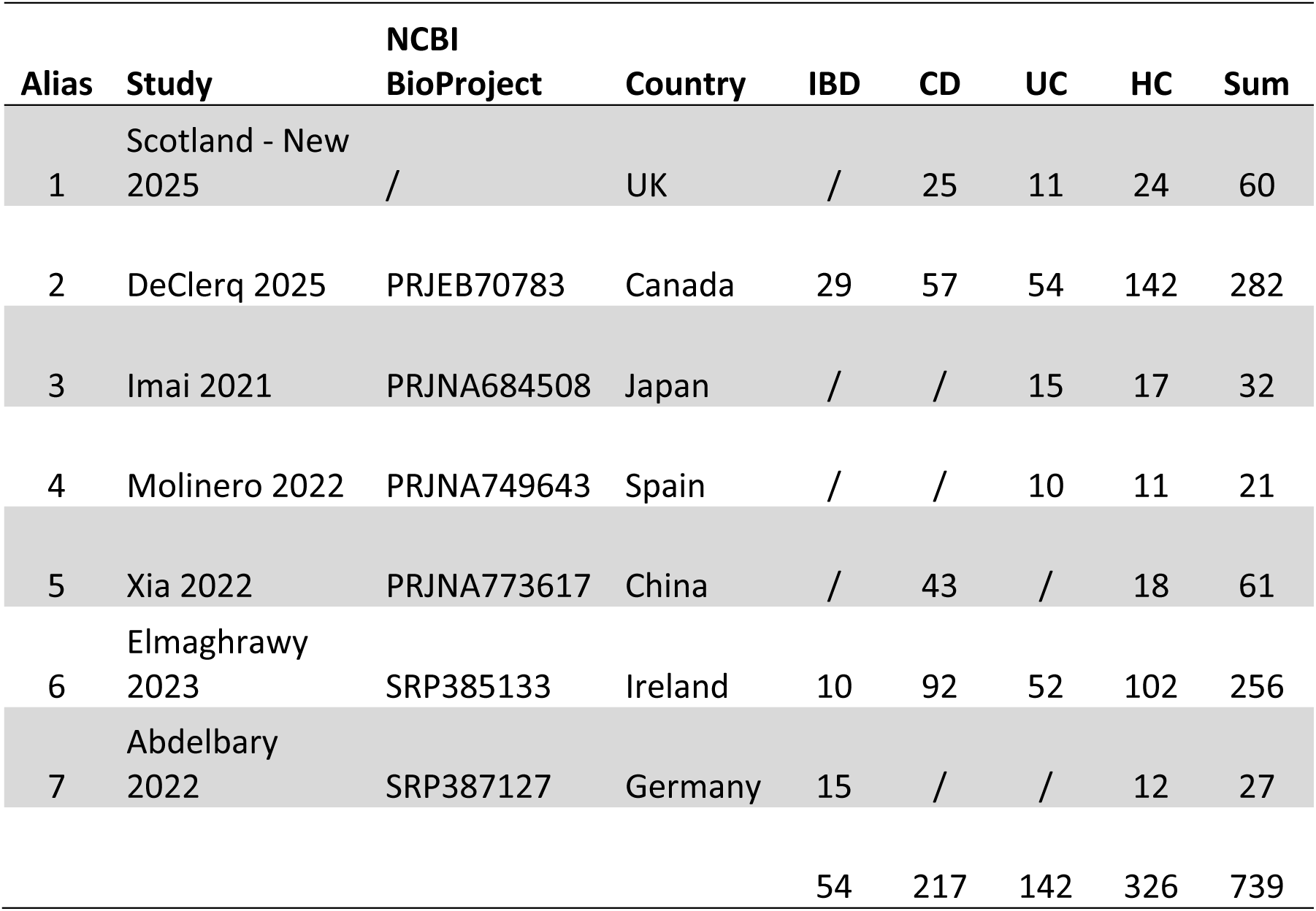
Characteristics of studies used in secondary 16S rRNA sequencing analysis.

### Oral microbial diversity is consistently reduced in IBD with greater disruption in CD

Primary random effects meta-analysis of the standardised mean difference in salivary Shannon diversity comparing 1136 IBD patients with 759 controls show a significant reduction in diversity in patients with IBD (standardised mean difference [95% CI]:-0.31 [-0.53,-0.08]) (Figs. S3 and S4). When examining IBD subtypes, there was significantly reduced alpha-diversity in patients with CD (standardised mean difference [95% CI]:-0.22 [-0.41,-0.04] but not in UC (standardised mean difference [95% CI]:-0.23 [-0.51, 0.04]) (Figs. S5 and S6, respectively). This difference was present during high disease activity and lost when considering low disease activity (Fig. S7).

To validate these findings within the secondary harmonised workflow, we next examined diversity metrics across the pooled datasets. We found that oral microbiota Shannon diversity and richness were lower in IBD compared to HC (Hedges’ SMD =-0.372, p < 0.001 and Hedges’ SMD =-0.387, p < 0.05, respectively) (Figs. 2A and B). These differences were driven mostly by CD for Shannon diversity (Hedges’ SMD =-0.442, p < 0.001), but not for richness (Hedges’ SMD =-0.450, p = 0.11, respectively) (Figs. 2A and B). UC also exhibited lower Shannon diversity but not richness (Hedges’ SMD =-0.205, p < 0.05 and richness; Hedges’ SMD =-0.108, p = 0.31, respectively) (Figs. 2A and B). There was no difference for these metrics between CD and UC. Pooled data analysis showed that the oral microbiota alpha-diversity was lower in IBD and CD compared to control, but not UC (Fig. 2B), with CD alpha-diversity significantly lower compared to UC (Fig. 2B). Beta-diversity analysis revealed global differences between IBD and IBD-subtypes from HC (all comparisons p < 0.001, IBD vs HC; *R^2^* = 0.007, IBD-subtype vs HC; *R^2^* = 0.012) (Figs. 2C and D). CD exhibited greater divergence from HC than UC (Fig. 2D). Additionally, differences in community composition were observed across geographical locations (Figs. S8A and B).

**Figure 2.**
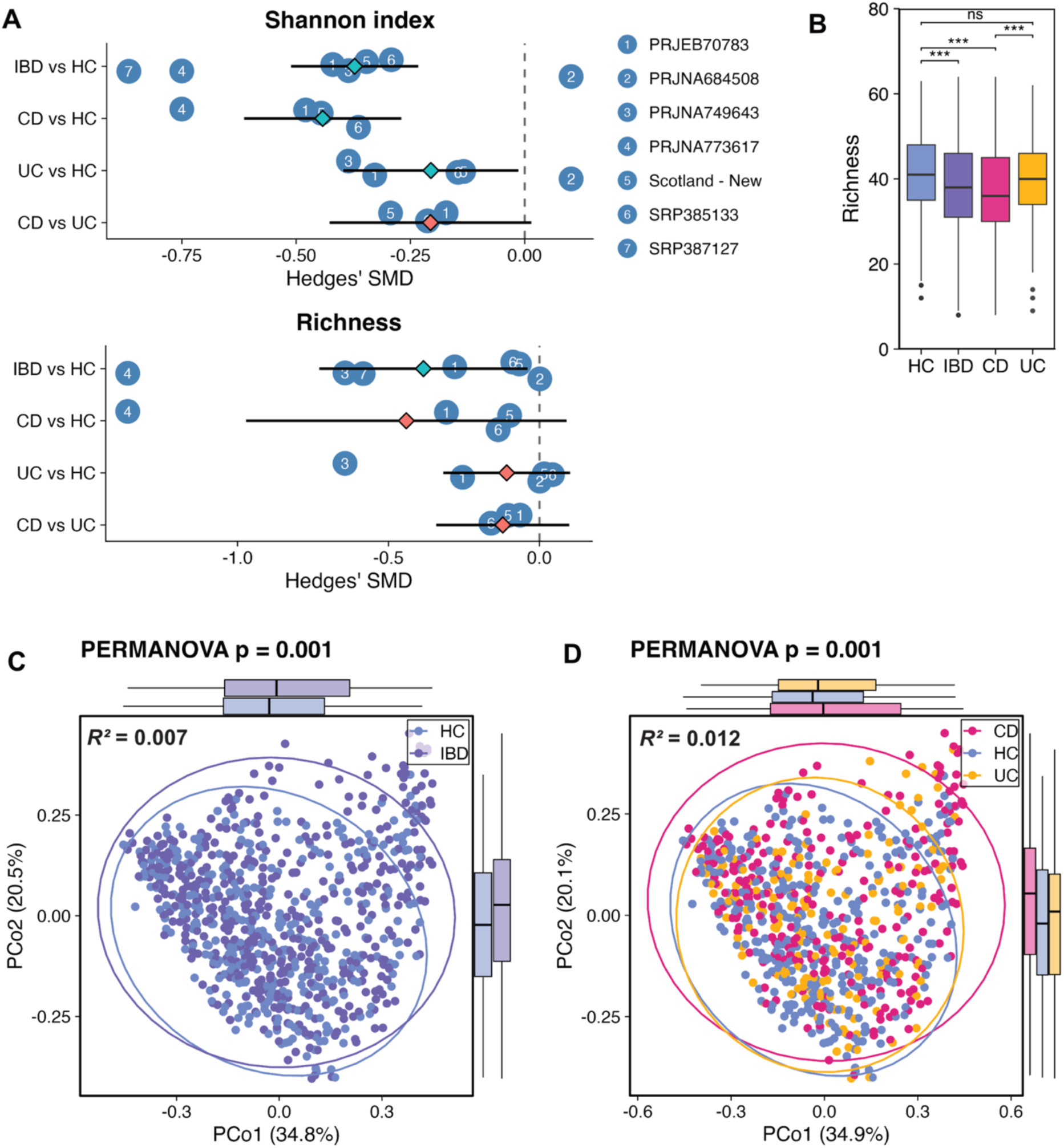
Diversity analysis between disease groups (A) Richness alpha diversity analysis on unrarefied genus–level between groups. Boxplots represent median with upper and lower boundaries representing 25^th^ and 75^th^ percentiles. Student’s t test, *p ≤ 0.05, **p ≤ 0.01, ***p ≤ 0.001, ns = non-significant. (B) Meta-analysis of the Shannon index and richness using Hedges’ SMD. Blue diamonds illustrate statistically significant difference (p < 0.05) and red diamonds illustrate non-significance. Lines represent upper and lower 95% confidence intervals. (C) Bray-Curtis principal coordinates analysis from IBD and HC patients and PERMANOVA analysis. Colours indicate disease group and ellipses represent 95% confidence bounds around group centroids. (D) Bray-Curtis principal coordinates analysis from CD, UC, and HC patients and PERMANOVA analysis. Colours indicate disease group and ellipses represent 95% confidence bounds around group centroids. All group comparisons were significant (p = 0.001).

### *Corynebacterium* integrates into core oral microbial network hubs specifically in IBD

As individual bacterial taxa act within structured communities rather than isolation, we next asked whether IBD altered the architecture of oral microbial interaction networks. To identify specific genera associated with IBD and IBD-subtypes, we performed a multivariate modelling on all microbial features using MaASLin2. Focusing first on IBD overall, the genera most strongly associated were *Corynebacterium* (coefficient = 0.57, q = 0.040), *Serratia* (coefficient = 0.52, q < 0.001) and *Streptoccocus* (coefficient = 0.16, q = 0.047) (Fig. 3A). Those mostly associated with HC were *Anaerovoracaceae.G1* (coefficient =-1.33, q < 0.001), *Porphyromonas* (coefficient =-1.31, q < 0.001), and *Ruminococcaceae.G1* (coefficient =-1.28, q < 0.001). Furthermore, our analysis revealed that *Arachnia* (coefficient = 0.58, q = 0.039) and *Serratia* (coefficient = 0.49, q = 0.017) were associated with CD (Fig. S9A). The genera mostly associated with HC were *Porphyromonas* (coefficient =-1.60, q < 0.001), *Hoylesella* (coefficient =-1.56, q < 0.001), and *Anaerovoracaceae.G1* (coefficient =-1.52, q < 0.001). Comparing differentially abundant microbial genera between UC and HC, we found that the genus *Serratia* (coefficient = 0.59, q = 0.014) was mostly associated with UC whilst those mostly associated with HC were *Anaerovoracaceae.G1* (coefficient =-1.10, q = 0.019), *Ruminococcaceae.G1* (coefficient =-1.03, q = 0.004), and *Porphyromonas* (coefficient =-0.84, q = 0.032) (Fig. S9B).

**Figure 3.**
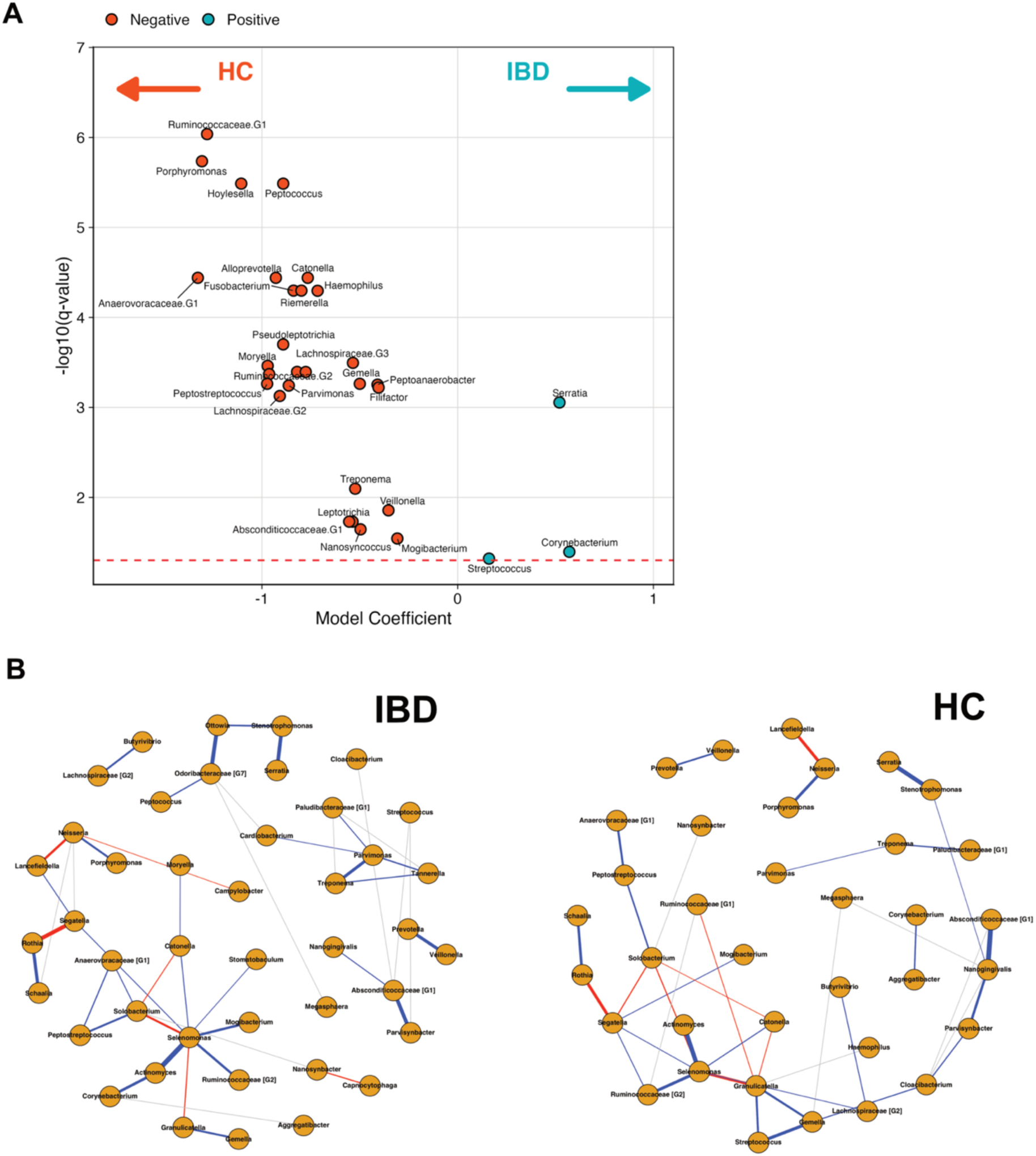
Altered taxonomic profiles and microbial network connectivity in IBD (A) Volcano plot displaying MaAsLin2 coefficients (x-axis) of differentially abundant genera using HC as the reference group. Positive coefficients indicate the genus is associated with higher levels in IBD (blue), while negative values indicate higher levels in HC (red). −Log10-transformed q values are shown on the y axis (Benjamini-Hochberg method). Plot shows only significantly associated taxa (q < 0.05). (B) Network analysis of microbial taxa for IBD and HC groups. Positive edges between nodes are indicated in blue, while negative edges between nodes are indicated in red. Thickness of edges indicate edge weight. Only significant interactions are shown.

Next, we applied the SPIEC-EASI framework to infer microbial interaction networks while accounting for the compositional and sparse nature of microbiome data. Both IBD and healthy control networks were organised around a central *Selenomonas* hub (Fig. 3B). *Selenomonas* is a known pathobiont associated with biofilm organisation and virulence in early childhood caries and periodontitis.^42^ In IBD, however, *Corynebacterium* was incorporated into this hub via *Actinomyces*, a configuration absent in health. Given the aerotolerant nature of *Corynebacterium*, its integration suggests a shift toward an inflammation-adapted, oxygen-tolerant oral community structure, analogous to the ‘oxygen-bloom’ community shifts described in the inflamed gut.^43,44^ Such network rewiring implies a shift in the ecological rules governing the oral microbiome rather than the expansion of a single pathogen. Furthermore, since *Corynebacterium* is primarily associated with sub and supragingival plaque, this might be indicative of increased plaque burden within IBD (REFs).

### Functional prediction implicates depleted butyrate production and elevated aromatic amino acid metabolism in CD

To determine whether these structural changes were accompanied by functional alterations, we used PICRUSt2 to predict MetaCyc pathway abundances. In CD, enriched pathways were predominately related to aromatic amino acid–derived and related metabolites degradation pathways (11/26 pathways), followed by l- arginine metabolism (3/26), cobalamin (vitamin B12) biosynthesis (3/26), carbohydrate acid degradation (3/26), cell wall/antimicrobial resistance functions (2/26) (Fig. 4A). In contrast, butyrate and short-chain fatty acid (SCFA) production pathways were enriched in HC (3/7 pathways). For UC, peptidoglycan biosynthesis was the most enriched pathway, whereas butyrate production was again mostly strongly associated with HC (Fig. 4B).

**Figure 4.**
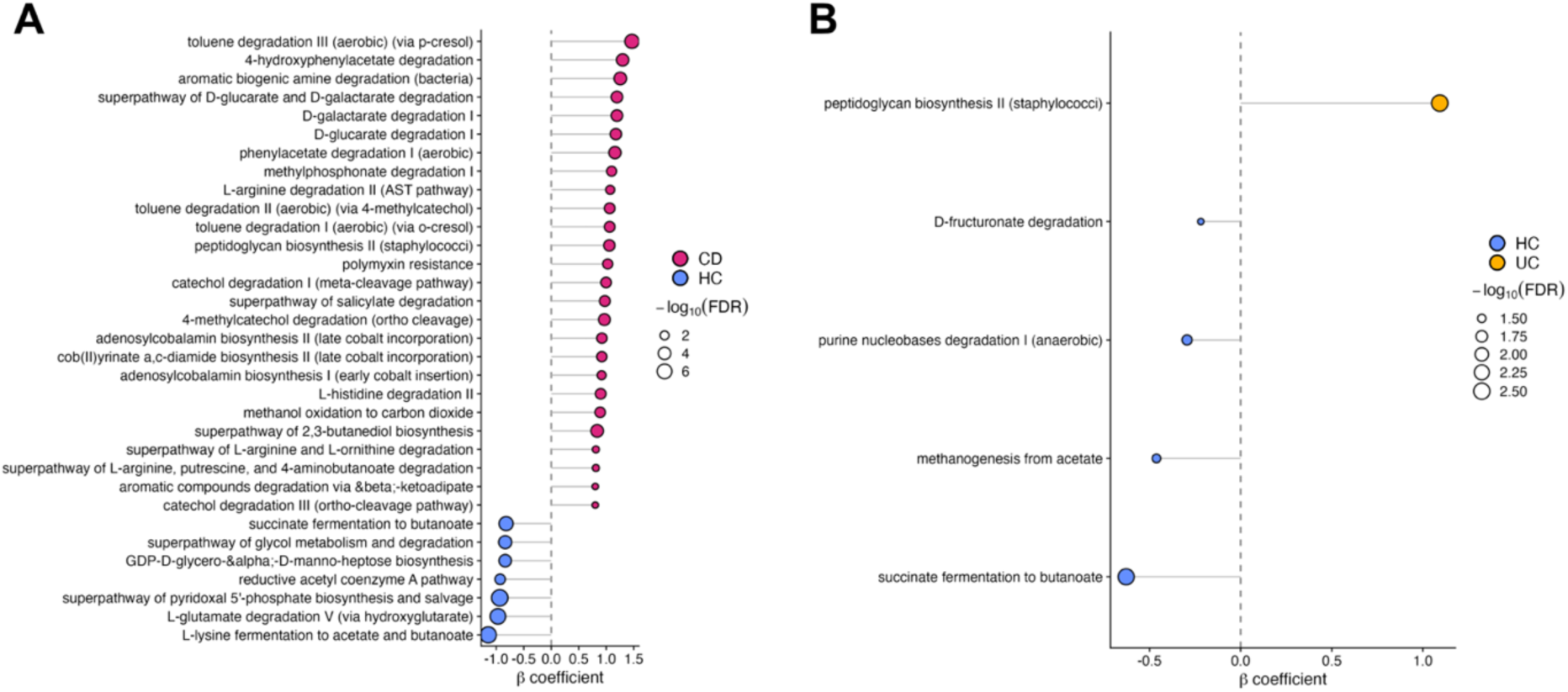
Functional imputation of the oral microbiota. MaAsLin2 coefficients (x-axis) of differentially abundant MetaCyc pathways imputed by PICRUSt2 using. (A) Positive coefficients indicate the pathway is associated with higher levels in CD (red), while negative values indicate higher levels in HC (blue). Pathways with MaAsLin coefficient > 0.8 or < –0.8 are shown. (B) Positive coefficients indicate the pathway is associated with higher levels in UC (yellow), while negative values indicate higher levels in HC (blue). −Log10-transformed q values are shown on the y-axis (Benjamini-Hochberg method).

Plot shows only significantly associated taxa (q < 0.05). Size of circle represents magnitude of −Log_10_-transformed q value.

These predicted functional changes suggest that the oral microbiome in CD shifts from a stable, SCFA-producing community towards a more inflammation-adapted metabolic state, characterised by increased amino acid utilisation and pathways linked to host immune signalling. Although imputed rather than directly measured, these predicted functions align with the observed structural changes and suggest that the oral microbiome in CD shifts towards an inflammation-adapted metabolic state, mirroring features of the inflamed gut microbiome.

### Oral microbiota-based machine learning achieves reproducible cross-cohort IBD classification

To test whether reproducible, cross-cohort oral microbiome signatures could support disease classification, we employed three machine learning cross-validation strategies to the oral microbiota datasets: within study cross validation was repeated 20 times with training and testing performed on the same study dataset; across study validation with classifiers trained on one study and tested on each other study to produce a study-study performance matrix; and leave-one-dataset-out (LODO) where trained models on all but one study and tested on the held-out dataset which was subsequently repeated for each study. We applied these strategies to train random forest models to classify IBD vs HC. We found that the LODO cross-validation resulted in one of the highest and most stable AUC score within the strategies (mean AUC score of 0.67, range 0.56–0.84) (Fig. 5A). Although this was the highest AUC score, classification accuracy of this strategy was limited. For CD, we found that the across study validation resulted in the highest and most stable AUC score within the strategies (mean AUC score of 0.63, range 0.50–0.83) (Fig. 5B). For UC, we found that across study validation resulted in the highest AUC score (mean AUC score of 0.55, range 0.41–0.72) (Fig. 5C). These results highlight the potential of using oral microbiota-based machine learning as screening tool for IBD classification when trained on diverse datasets, however, higher taxonomic resolution of the microbial features may be needed to improve accuracy.

**Figure 5.**
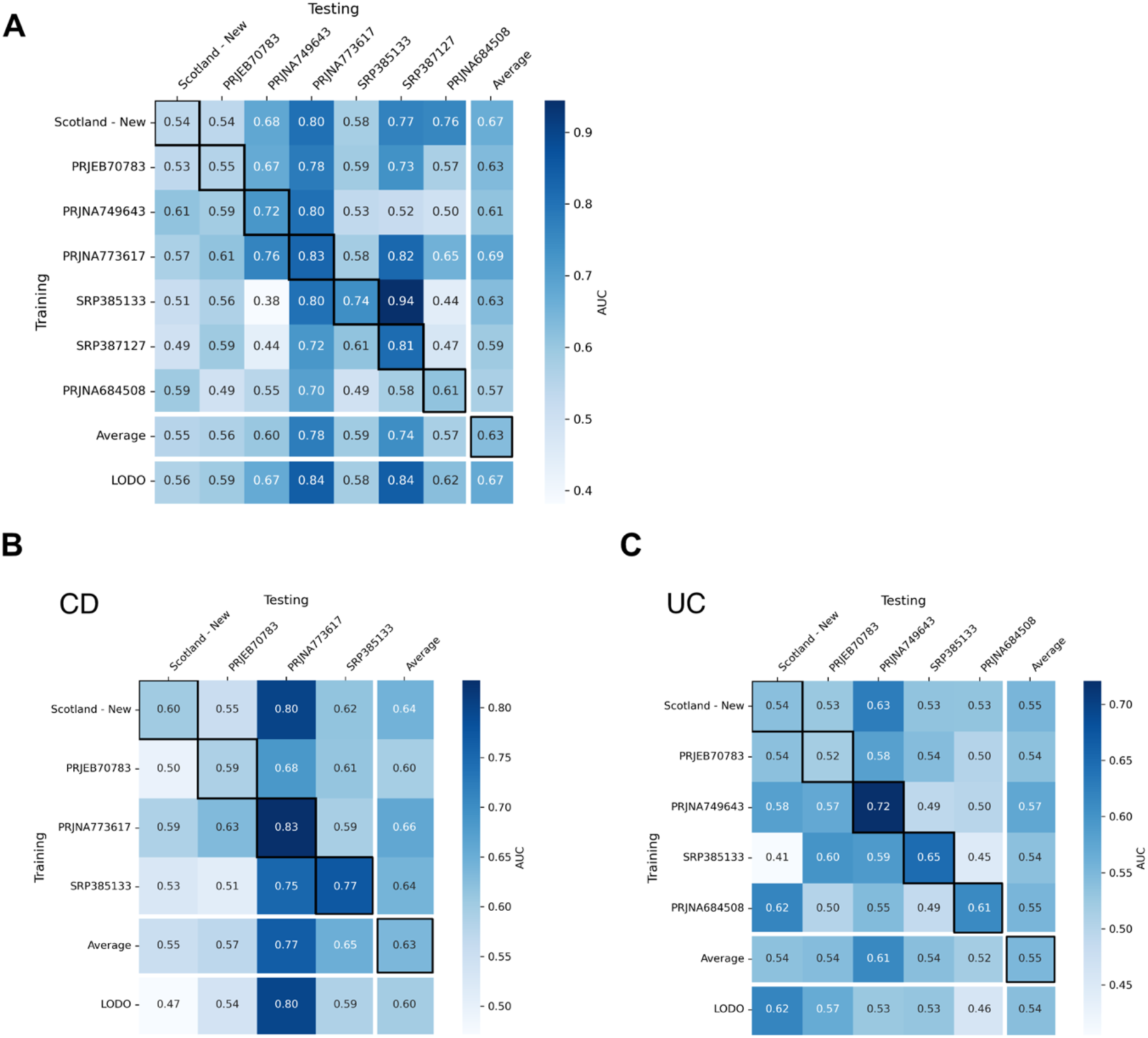
Classification accuracy of IBD and IBD subtypes using oral microbiota features. Heatmaps illustrating AUC from random forest classifier matrices. Diagonals represent within models and off diagonals represent across study model comparisons. LODO comparison represent training on all studies, leaving one study out, and testing on the left-out study (column). (A) represents IBD, (B) represents CD, and (C) represents UC.

## Discussion

The human oral microbiome has increasingly been implicated in IBD, yet findings across studies have remained fragmented due to heterogeneity in sampling, study design and analytical approaches. By integrating oral microbiome datasets across multiple independent cohorts within a unified analytical framework, we demonstrate that IBD particularly CD, is characterised not simply by compositional change, but by reproducible ecological restructuring of the oral microbial community. This restructuring is most evident at the level of community organisation, where consistent alterations in diversity, taxonomic composition and microbial interactions converge to define a CD-dominant ecological signature distinct from both ulcerative UC and health. These findings suggest that the oral microbiome in CD is governed by altered ecological rules, rather than isolated taxonomic shifts, providing a unifying systems-level framework for the oral–gut axis in IBD.^3–5,45^

Emerging evidence points to a role for the oral-gut axis in IBD pathogenesis. First, oral bacteria can directly drive the Th1 and Th17 effector responses central to CD pathogenesis in experimental models - saliva from patients with CD is colitogenic in germ-free hosts, and individual oral-origin strains such as *Klebsiella pneumoniae* 2H7 recapitulate colitis in susceptible mice.^17,18^ Second, the inflamed gut produces metabolic cues (nitrate, amino-acid catabolites) that create a niche for ectopic oral colonisation, making the oral–gut axis inherently feed-forward rather than one-way.^22^ Third, the oral mucosa itself is an inflammation-primed immune tissue, with a resident stromal–neutrophil programme and cell-specific expression of periodontitis susceptibility genes.^16^ Our data provides evidential data for this scaffold: a reproducible oral signature in IBD, structurally dominated by CD, detectable across geographies, age groups and sequencing platforms.

The most mechanistically informative signal emerged at the level of community organisation rather than individual taxa. To capture this, we inferred microbial interaction networks reflecting patterns of co-occurrence and exclusion, providing insight beyond relative abundance alone. In health, oral communities were organised around a central *Selenomonas* hub, consistent with its established role in oral biofilm structure. In IBD, however, this architecture was rewired, with *Corynebacterium* integrating into the central network via *Actinomyces*, a configuration not observed in health. Given the aerotolerant nature of *Corynebacterium*, this shift is consistent with an inflammation-adapted, oxygen-tolerant ecological state. This pattern mirrors the ‘oxygen bloom’ described in the inflamed gut, where obligate anaerobes are replaced by facultative and aerotolerant organisms ^3,43,44^. In this framework, CD represents the point at which this upstream ecological adaptation is most apparent. Such community-level reorganisation, rather than isolated taxonomic differences, is unlikely to be captured by single-taxon analyses and becomes evident primarily through harmonised, multi-cohort approaches.

Notably, *Corynebacterium* emerged as a central node within disease-associated networks. While this genus has not been prominently implicated in Crohn’s disease pathogenesis, it is a well-recognised component of oral biofilm communities and frequently co-occurs with structural taxa such as *Actinomyces*.^46,47^ Its apparent integration into network hubs in Crohn’s disease is therefore unlikely to reflect a direct pathogenic role, but rather a shift in the underlying ecological organisation of the oral microbiome. In this context, *Corynebacterium* may serve as a marker of altered community architecture, potentially reflecting changes in oxygen tolerance, biofilm structure, or host inflammatory milieu. Importantly, this highlights that network-derived centrality does not equate to causality but instead provides insight into system-level reorganisation that is not captured by differential abundance analyses alone.

Functional pathway imputation using PICRUSt2 reinforced this interpretation. In CD, depletion of butyrate and short-chain fatty acid production pathways was paired with enrichment of aromatic amino-acid–derived metabolite degradation and L-arginine metabolism. Of relevance, L-arginine metabolism feeds nitric oxide signalling and Th17-associated gut inflammation,^48^ while aromatic amino acid catabolism and the loss of butyrate both indicate a move away from immunoregulatory, carbohydrate-based metabolism toward a stressed microbial state associated with impaired mucosal tolerance.^49^ Recent evidence shows that inflammation-associated nitrate drives ectopic colonisation by oral *Veillonella parvula*,^22^ the predicted metabolic shift is similarly expected if the oral microbiome were being restructured along oralisation-permissive lines. Although imputed rather than directly measured, these predicted functions align closely with the observed structural changes and with established immunometabolic pathways in gut inflammation.

Multivariate modelling identified a reproducible set of genera associated with IBD across independent cohorts. *Corynebacterium*, *Serratia* and *Streptococcus* were consistently enriched in IBD, while *Porphyromonas*, *Anaerovoracaceae* G1 and *Ruminococcaceae* G1 were enriched in health. These associations were robust to multivariate adjustment for geography, age group and hypervariable region sequenced. Notably, the most extensively studied oral IBD-associated taxa in the mechanistic literature: *Fusobacterium nucleatum*, *Campylobacter concisus* and *Porphyromonas gingivalis* were not prominent in our analysis. This likely reflects the dominance of salivary sampling in available datasets, with a near-complete absence of dental plaque studies. These canonical taxa are primarily biofilm organisms, most reliably recovered from subgingival plaque, and their absence in a saliva-dominated synthesis is expected rather than contradictory.^46,50^

Machine-learning classifiers trained on harmonised oral microbiota genus features achieved limited performance to discriminate IBD and CD across cohorts (mean AUC ∼ 0.67 for IBD, 0.63 for CD in the best-performing cross-validation strategies), robust to leave-one-dataset-out and across-study validation. These values are not yet clinically actionable on their own, but they establish an important point: a reproducible, learnable oral signal exists across independent studies, geographies and sequencing regions. Further gains are most likely to come from strain-level taxonomic resolution and functional profiles from shotgun metagenomics, and from paired dental plaque sampling, rather than from additional saliva 16S rRNA data alone.

Limitations: The near-absence of dental plaque studies is a key data gap in the field. Saliva, while accessible, is a composite sample that dilutes and potentially obscures biofilm-associated signals from the subgingival and supragingival plaque, the niches most likely to harbour canonical IBD-associated pathobionts and to interface directly with mucosal immune surveillance.^50^ Short-read 16S rRNA sequencing restricts resolution to the genus level in most cases; species-and strain-level variation, achievable via long-read sequencing or shotgun metagenomics, may unmask ecologically and clinically relevant structure invisible here.^51^ The analysis is cross-sectional; longitudinal and interventional data are needed to distinguish cause from consequence and to track oralisation events in vivo. Differences between IBD cohorts and geographic heterogeneity (Figs. S8A and B) may account in part for subtype differences and underscore that single-centre oral microbiome studies risk confounding by local environmental factors; harmonised multi-cohort approaches of the kind used here are essential.

In conclusion, these findings demonstrate that the oral microbiome in IBD is characterised not only by reduced diversity but by reproducible, cross-cohort restructuring of microbial community organisation, most prominently in CD. This reorganisation is evident at the level of ecological networks, where altered patterns of microbial interaction define a disease-associated community state, including, but not limited to the integration of taxa such as *Corynebacterium* into core network architecture. By shifting the focus from individual taxa to system-level behaviour, these data provide a unifying framework for understanding the oral-gut axis in IBD. This framework highlights the need for next-generation studies that integrate strain-resolved microbiomics, functional profiling and longitudinal sampling to elucidate how oral microbial ecology contributes to intestinal inflammation and disease progression.

## Ethics declarations

Ethical approval for Mitochondrial DAMPs as mechanistic biomarkers of mucosal inflammation in Crohn’s Disease (MUSIC study; www.musicstudy.uk; ClinicalTrials.gov NCT04760964) was obtained from East Scotland Ethics Committee (REC 19/ES/0087). The study was conducted in accordance with the UK Policy Framework for Health and Social Care Research, and all participants provided informed consent.

## Data availability statement

The Scotland – New sequencing data generated in this study have been deposited in the Sequence Read Archive under accession number PRJNA1458073.

## Author Contributions

Author contributions are listed according to the CRediT taxonomy: conceptualisation: R.J.W., D.I.F.W., G.T.H.; data curation: R.J.W., D.I.F.W., G.M., J.I.; formal analysis: R.J.W., D.I.F.W.; investigation: R.J.W., D.I.F.W., G.T.H.; methodology: R.J.W., D.I.F.W.; supervision: G.T.H.; writing—original draft: R.J.W., D.I.F., G.T.H.; writing—review and editing: all authors reviewed the manuscript.

## Data Availability

The Scotland - New sequencing data generated in this study have been deposited in the Sequence Read Archive under accession number PRJNA1458073 and will be available upon publication.

