## Supplementary material for "Reproducible ecological restructuring of the oral microbiome defines the oral-gut axis in Crohn’s disease"

**Supplementary data**

### Tables

| REF | STUDY | SAMPLE TYPE | PATIENT GROUP | AGE DISTRIBUTION (YEARS) | TYPE OF CONTROL | SAMPLE NUMBERS INCLUDED IN MICROBIOME ANALYSIS | | | | SELECTED CONFOUNDING VARIABLES | | | | | |
|---|---|---|---|---|---|---|---|---|---|---|---|---|---|---|---|
| | | | | | | Total IBD | UC | CD | Control | Treatment naive | Antibiotic exposure >1 month | Dental examination performed | Patient fasted before sample? | Was saliva sample unstimulated? | Was dietary data available? |
| **51** | Elmaghrawy et al, 2022, Europe | Saliva - tongue swab | Paediatric | 4.06 - 16.69 | Symptomatic Non IBD and Healthy control | 146 | 52 | 94 | 102 | Yes | No | Yes | Yes | Yes | No |
| **52** | Abdelbary et al, 2022, Europe | Saliva, Stool | Adult | 35.6 (mean) | Healthy control | 14 | - | - | 12 | No | No | No | Unclear | Unclear | No |
| **53** | Shin et al, 2022, Asia | Saliva, Stool, Ileal biopsy | Adult | 35.7 ± 11.2 | No control | 30 | - | 30 | 0 | No | No | No | Unclear | Unclear | No |
| **54** | Somineni et al, 2021, North America | Saliva - tongue swab, Subgingival plaque, Buccal mucosa, Stool | Paediatric | 5.1 - 19.3 | Healthy control | 47 | 17 | 30 | 18 | Some - 55% | No | Yes | Unclear | Unclear | No |
| **55** | Elzayat et al, 2023, Asia | Saliva | Adult | 16 - 52 | Healthy control | 40 | - | - | 40 | Some - 15% | No | Yes | Yes | Yes | No |
| **56** | Räisänen et al, 2023, Europe | Saliva | Paediatric | 11.9 ± 1.6 | Healthy control | 14 | - | - | 40 | Yes | No | No | Unclear | Yes | No |
| **57** | Imai et al, 2021, Asia | Saliva, Stool | Adult | 16 - 39 | Healthy control | 58 | 42 | 16 | 45 | No | No | Yes | Unclear | Unclear | No |
| **58** | Kang et al, 2023, Asia | Saliva | Adult | 38.3 ± 14.17 | Healthy control | 398 | 175 | 127 | 100 | No | No | No | Unclear | Unclear | No |
| **59** | Hu et al, 2021, Asia | Saliva, Stool | Adult | 21 - 67 | Healthy control | 25 | - | - | 25 | No | No | Yes | Yes | Yes | No |
| **60** | Goel et al, 2023, Europe | Saliva | Adult | 16 - 79 | Healthy control | 123 | - | 123 | 45 | No | No | Yes | Yes | Yes | No |
| **61** | Sohn et al, 2023, North America | Saliva, Subgingival plaque, Stool | Adult | 53.48 (mean) | Healthy control | 25 | - | 25 | 25 | No | Unclear | Yes | Unclear | Unclear | Yes |

| | | | | | | | | | | | | | | | |
|---|---|---|---|---|---|---|---|---|---|---|---|---|---|---|---|
| **62** | Xu et al, 2023, Asia | Saliva, Buccal swab, Stool | Adult | 44.82 ± 9.91 | Healthy control | 40 | 40 | - | 21 | No | No | Yes | No | No | No |
| **63** | Zhang et al, 2020, Asia | Saliva | Adult | Not stated | Healthy control | 60 | - | 60 | 31 | No | No | No | Unclear | Unclear | No |
| **64** | Qi et al, 2021, Asia | Saliva | Adult | 27.6 ± 6.2 | Healthy control | 22 | 10 | 12 | 8 | No | No | Yes | Yes | No | No |
| **65** | Kelsen et al, 2015, North America | Subgingival plaque | Paediatric | 2 - 21 | Healthy control | 79 | - | 79 | 74 | No | Yes | No | Yes | N/A | No |
| **66** | Xia et al, 2022, Europe | Saliva - tongue swab, Sputum, Ileal stoma swab | Adult | 33.2 ± 11.6 | Healthy control | 43 | - | 43 | 18 | No | No | No | Unclear | Yes | No |
| **67** | Said et al, 2014, Asia | Saliva | Adult | 36.9 ± 14.5 | Healthy control | 35 | 14 | 21 | 24 | No | Unclear | No | Unclear | Yes | No |
| **68** | Molinero et al, 2022, Europe | Saliva | Adult | 18 - 44 | Healthy control | 10 | 10 | - | 11 | No | Unclear | No | Yes | Yes | No |
| **69** | Hu et al, 2022, Asia | Saliva | Adult | 21 - 67 | Healthy control | 41 | - | 41 | 24 | No | No | Yes | Yes | Yes | No |
| **70** | Monleon-Getino et al, 2023, Europe | Saliva, Stool | Paediatric | 9 - 16 | Healthy control | 24 | 11 | 13 | 8 | Yes | No | No | Unclear | Yes | No |
| **71** | Xun et al, 2018, Asia | Saliva | Adults | 43.8 ± 13.9 | Healthy control | 79 | 54 | 13 | 25 | No | No | Yes | Unclear | Yes | No |
| **72** | Park et al, 2022, Asia | Saliva, Stool, Urine, Serum | Adult | 32 (mean) | Healthy control | 19 | - | - | 19 | No | No | No | Yes | Yes | No |
| **73** | DeClercq et al, 2025, North America | Saliva | Adult | 55.6 (36-69) | Healthy control | 160 | 65 | 64 | 160 | No | No | No | No | Yes | Yes |
| **74** | Sun et al, 2024, Asia | Saliva | Adult | 36 (20-45) | Healthy control | 10 | 10 | 0 | 12 | Yes | Yes | Yes | Unclear | Yes | Yes |
| **75** | Xu et al, 2025, Asia | Buccal Swab | Adult | Not stated | Healthy control | 53 | 53 | 0 | 28 | No | No | Yes | No | Yes | NA |

**Table S1.** Summary of the characteristics of each included study, including patient demographics, number of microbiome samples analysed, and selected confounding variables considered in the study design.

| | STUDY | SEQUENCING PLATFORM | SEQUENCING METHOD | HYPERVARIABLE REGION | SAMPLE COLLECTION KIT | SAMPLE STORAGE | TRANSPORT METHOD | DNA EXTRACTION METHOD | BIOINFORMATIC TOOLS | REFERENCE DATABASE | CORRECTION FOR MULTIPLE TESTING | PICRUSt | PREDICTION MODEL |
|---|---|---|---|---|---|---|---|---|---|---|---|---|---|
| 51 | Elmaghrawy et al, 2022 | Illumina MiSeq | 16S rRNA | V1-V2 | Polyurethane sponge swab [CultureSwab EZ, Becton, Dickinson] | Immediately frozen at -80°C | Not stated | MasterPure Complete DNA/RNA Purification Kit [Epicentre Biotechnologies ] | Dada2, Phyloseq, Microbiome, MicroViz, MaAsLin 2, DeSeq2, Siamcat, mothur, | eHOMD | Yes (Benjamini–Hochberg) | Yes | Yes |
| 52 | Abdelbary et al, 2022 | Illumina MiSeq | 16S rRNA | V3-V4 | Sterile 100 ml container (Sarstedt) | Immediately frozen at -72°C | Saliva aliquots transferred to DNA stabilizer tubes (DNA/ RNA Shield Lysis Tubes-Microbe, Cat #. R1103, Zymo Research) | QIAamp DNA Mini Kit (Qiagen) | UPARSE IMNGS, FastTree, Rhea, Microeco, Vegan, BLAST | Not specified | Yes (Benjamini–Hochberg) | Yes | No |
| 53 | Shin et al, 2022 | Illumina MiSeq | 16S rRNA | V3-V4 | Saliva collection kit (Cat. PDX-026; PDXen Biosystems Co.) | Stored at room temperature until extraction | Transported at room temperature (15–30 ◦C) | QIAamp DNA Microbiome Kit (Qiagen) | QIIME2, DADA2, MAFFT, FasTtree2, q2-feature-classifier | SILVA | Not stated | No | No |
| 54 | Somineni et al, 2021 | Illumina MiSeq | 16S rRNA | V4 | DNA swabs (Isohelix) | Immediately frozen at -80°C | Not stated | Biostic Bacteremia DNA Isolation Kit (MO BIO Laboratories Inc.) | Pipeline not described, metagenomeSeq | SILVA | Yes false discovery rate (FDR) method | No | Yes |
| 55 | Elzayat et al, 2023 | Oxford Nanopore sequencer | 16S rRNA | Full length | Sterile container, 1ml centrifuged to pellet the cells | Not stated | Not stated | Wizard HMW DNA extraction kit, nanodrop (Termo Scientifc) | Kraken taxonomic sequence classifcation system, Partek® Genomics Suite® software, Microbiome Analyst 2.0 platform | Not stated | Not stated | No | No |
| 56 | Räisänen et al, 2023 | Illumina MiSeq | 16S rRNA | V3-V4 | Oragene® DNA Self-Collection Kit (DNA Genotek) | Stored at room temperature until extraction | Not stated | In house - TS-tailed-1S dual index protocol | CLC Genomics Workbench, adonis, vegan, DESeq2, Phyloseq, Microbiome, STAMP | SILVA | Yes (Benjamini–Hochberg) | Yes | No |
| 57 | Imai et al, 2021 | Illumina MiSeq | 16S rRNA | V4 | Not stated | Not stated | Not stated | DNeasy Blood & Tissue Kit (Qiagen) | DADA2, mothur | Ribosomal Database Project | Not stated | No | No |
| 58 | Kang et al, 2023 | Illumina MiSeq | 16S rRNA | V3-V4 | Saliva collection kit (Cat. PDX-026; PDXen Biosystems Co.) | Stored at room temperature until extraction | Not stated | QIAamp DNA Microbiome Kit (Qiagen) | FASTP, Kraken2, phyloseq, microbiome, mothur, mixOmics | SILVA | Not stated | No | Yes |
| 59 | Hu et al, 2021 | Illumina HiSeq 4000 | Metagenomic | N/A | OMNIgene Discover Kit 505 (DNA Genotek) | Stored at room temperature until extraction at 4 weeks | Not stated | Exgene Clinic SV Mini kits (GeneAll Biotechnology) | Skewer, BWAMEM, MetaPhlAn2, StrainPhlAn, Vegan, HUMAnN2 | hg19 human reference genome | Yes (Benjamini–Hochberg) | N/A | No |

| | | | | | | | | | | | | | |
|---|---|---|---|---|---|---|---|---|---|---|---|---|---|
| 60 | Goel et al, 2023 | Roche 454 GS-FLX Titanium sequencer. | 16S rRNA | V1-V3 | Universal container | Frozen at -70°C within 3 hours | On ice | Genelute DNA extraction kit (Sigma-Aldrich) | mothur, AmpliconNoise algorithm, Uchime, mothur2oligo, BLAST | SILVA eHOMD | Yes (Bonferroni correction) | No | No |
| 61 | Sohn et al, 2023 | Illumina MiSeq | 16S rRNA | V3-V4 | 15 mL tube | Not stated | On ice | QIAamp DNA Mini kit (Qiagen) | QIIME2, DADA2, ANCOM | Genome taxonomy database | Yes (Dunn's multiple comparison) | Yes | No |
| 62 | Xu et al, 2023 | Illumina MiSeq | 16S rRNA | V3-V4 | Sterile DNase-free and RNase-free centrifuge tube | Not stated | Not stated | Not clear | Vsearch, Usearch, STAMP, Pheatmap, Vegan | Ribosomal Database Project | Not stated | **Yes** | No |
| 63 | Zhang et al, 2020 | Illumina MiSeq | 16S rRNA | V3-V4 | Sterile container | Frozen at -70°C within 3 hours | On ice | QIAamp DNA Mini kit (Qiagen) | QIIME, UPARSE, Usearch | Ribosomal Database Project | Not stated | Yes | No |
| 64 | Qi et al, 2021 | Illumina MiSeq | 16S rRNA | V3-V4 | Not stated | Preserved in freezing medium and stored at −80 °C within 2 hours | Ice bag and polystyrene foam container | Bacterial DNA kit (Omega Bio-Tek) | QIIME2, DADA2 | SILVA | Unclear | **Yes** | Yes |
| 65 | Kelsen et al, 2015 | 454 Life Sciences FLX instrument | 16S rRNA | V1-V2 | Sterile microbrush | Immediately frozen at -80°C | Not stated | PSP Spin Stool DNA Plus Kit (STRATEC Molecular) | QIIME, UCLUST | Greengenes | Yes (Benjamini–Hochberg) | No | Yes |
| 66 | Xia et al, 2022 | Illumina MiSeq | 16S rRNA | V3-V4 | Sterile cotton swab and immediately placed into a 2-ml sterile container | Stored at −80 °C within 2 hours | Ice pack | E.Z.N.A.® soil DNA Kit (Omega Bio-tek) | Mothur, FLASH, UPARSE, UCHIME | SILVA | Yes (Benjamini–Hochberg) | Yes | No |
| 67 | Said et al, 2014 | 454 GS FLX Titanium; 454 GS JUNIOR | 16S rRNA | V1-V2 | Not stated | Immediately frozen at -80°C | Not stated | Morita et al 2007 method | UCLUST, BLAST, Vegan | Bespoke database | Yes (Benjamini–Hochberg) | No | No |
| 68 | Molinero et al, 2022 | Illumina MiSeq | 16S rRNA | V3-V4 | Not stated | Stored at −80 °C | Ice bath | MasterPure™ Complete DNA and RNA Purification Kit (Epicentre) | DADA2, phyloseq, vegan, microbiome maker, yingtools2 | SILVA | Yes (Benjamini–Hochberg, Holm's multiple comparison testing correction) | No | No |
| 69 | Hu et al, 2022 | Illumina HiSeq 4000 | Metagenomic | N/A | OMNIgene Discover Kit 505 [DNA Genotek] | Stored at room temperature until extraction at 4 weeks | Not stated | Exgene Clinic SV Mini kits [GeneAll Biotechnology] | Terra workspace, bioBakery meta'omics workflow, KneadData, MetaPhlan3, HUMAnN3, UniRef-based protein sequence, Vegan, MaAsLin2 | MetaPhlan3 | Yes false discovery rate (FDR) method | N/A | Yes |
| 70 | Monleon-Getino et al, 2023 | NovaSeq 6,000 | Metagenomic | N/A | ORAcollect (OC-175, DNA Genotek) | Stored at (−20 ± 5)°C | Not stated | QIAsymphony DSP Virus/ Pathogen Kit and QIAsymphony SP (QIAGEN) | GAIA, phyloseq, FAMLI, UniProt API, BDbiost3 | GAIA database | Yes (unclear method) | N/A | Yes |
| 71 | Xun et al, 2018 | Illumina MiSeq | 16S rRNA | V3-V4 | 50-mL sterile Eppendorf tube | Stored at −80 °C | Ice bag and polystyrene foam container | TIANamp Bacteria DNA Kit (Tiangen Biotech) | QIIME, FLASH, Cytoscape | Greengenes | Unclear | Yes | No |

| | | | | | | | | | | | | | |
|---|---|---|---|---|---|---|---|---|---|---|---|---|---|
| *72* | Park et al, 2022 | Illumina MiSeq | 16S rRNA | V3-V4 | 15-ml falcon tube | Unclear | Unclear | FastDNA Spin Kit for Soil (MP Biomedicals) | QIIME, EzBioCloud MTP pipeline, Phyloseq, vegan, CASPER, VSEARCH, UCLUST, BLAST | SILVA | Unclear | No | No |
| *73* | DeClercq et al, 2025 | Illumina MiSeq | 16S rRNA | V4-V5 | 50-mL sterile tube | Stored at −80 °C | On ice | QIAamp 96 PowerFecal QIAcube HT kit | QIIME 2, DADA2, scikit-learn, HOMD | eHOMD | Unclear | No | No |
| *74* | Sun et al, 2024 | Illumina MiSeq | 16S rRNA | V4 | 15-ml falcon tube | Stored at −80 °C | Ice pack | QIAamp DNA Mini kit | QIIME | SILVA | Unclear | No | No |
| *75* | Xu et al, 2025 | Unclear895z | 16S rRNA | V3-V4 | Oral swab | Stored at −80 °C | Unclear | E.Z.N.A. Tissue DNA kit | BLAST | SILVA | Unclear | Yes | Yes |

**Table S2:** Summary of the methodology and analysis of included studies

Table S3

| REF | STUDY | SELECTION | | | | COMPARIBILITY | EXPOSURE | | | Score |
|---|---|---|---|---|---|---|---|---|---|---|
| | | Q1 | Q2 | Q3 | Q4 | Q5 | Q6 | Q7 | Q8 | |
| 51 | Elmaghrawy et al, 2022 | * | * | - | - | ** | * | * | * | 7 |
| 52 | Abdelbary et al, 2022 | * | - | - | * | *- | * | - | - | 4 |
| 53 | Shin et al, 2022 | * | - | - | - | -- | * | * | - | 3 |
| 54 | Somineni et al, 2021 | * | - | * | - | *- | * | - | * | 5 |
| 55 | Elzayat et al, 2023 | * | - | - | * | *- | * | - | * | 5 |
| 56 | Räisänen et al, 2023 | - | - | * | - | *- | - | * | * | 4 |
| 57 | Imai et al, 2021 | * | - | - | * | -* | * | - | - | 4 |
| 58 | Kang et al, 2023 | * | - | - | * | *- | * | * | * | 6 |
| 59 | Hu et al, 2021 | * | - | - | - | ** | * | - | - | 4 |
| 60 | Goel et al, 2023 | * | - | - | * | ** | * | * | - | 6 |
| 61 | Sohn et al, 2023 | * | - | * | * | ** | * | * | * | 8 |
| 62 | Xu et al, 2023 | * | - | - | * | *- | * | * | * | 6 |

| | | | | | | | | | | |
|---|---|---|---|---|---|---|---|---|---|---|
| 63 | Zhang et al, 2020 | * | - | - | * | *- | * | * | * | 7 |
| 64 | Qi et al, 2021 | * | - | - | - | ** | * | * | * | 6 |
| 65 | Kelsen et al, 2015 | * | - | * | * | *- | * | * | * | 7 |
| 66 | Xia et al, 2022 | * | - | - | * | *- | * | * | * | 6 |
| 67 | Said et al, 2014 | * | - | - | - | *- | - | * | * | 4 |
| 68 | Molinero et al, 2022 | * | - | - | * | ** | * | * | - | 6 |
| 69 | Hu et al, 2022 | * | - | * | * | ** | * | * | - | 7 |
| 70 | Monleon-Getino et al, 2023 | * | * | - | * | *- | * | * | - | 6 |
| 71 | Xun et al, 2018 | * | - | * | * | ** | * | * | - | 7 |
| 72 | Park et al, 2022 | * | - | * | * | *- | * | * | - | 6 |
| 73 | DeClercq et al, 2025 | - | - | * | * | -- | - | * | - | 3 |
| 74 | Sun et al, 2024 | * | - | - | * | ** | * | * | - | 6 |
| 75 | Xu et al, 2025 | * | - | * | * | -* | * | * | - | 6 |

**Table S3**. Risk of bias was assessed using the Newcastle–Ottawa Scale (NOS), which evaluates studies across three domains: selection of study groups (0–4 points), comparability of groups (0–2 points) utilising dental exam and use of antibiotics in preceding month as main confounding variables, and ascertainment of the outcome or exposure (0–3 points). A maximum of 9 points can be awarded, with higher scores indicating lower risk of bias.

| **Scotland – New dataset** | | | | | | | |
|---|---|---|---|---|---|---|---|
| | **CD** | | | **UC** | | **HC** | **p-value** |
| Numbers, n | 24 | | | 11 | | 24 | |
| Age, median ± IQR | 31 ± 9 | | | 59 ± 26 | | 36 ± 20 | 0.01 |
| Male:Female, n | 17:8 | | | 7:4 | | 9:15 | 0.08 |
| **Montreal Classification** | | | | | | | |
| | **n** | **%** | | **n** | **%** | | |
| *L1 Ileal* | 4 | 17 | *E1 Proctitis* | 0 | 0 | | - |
| *L2 Colonic* | 9 | 37 | *E2 Left-sided* | 2 | 18 | | - |
| *L3 Ileocolonic* | 11 | 46 | *E3 Pancolitis* | 9 | 82 | | - |
| *+/- L4 Upper GI* | 1 | | | | | | |
| *B1 Non-stricturing, non-penetrating* | 12 | 50 | *S0 Remission* | 0 | 0 | | - |
| *B2 Stricturing* | 9 | 38 | *S1 Mild* | 1 | 12 | | - |
| *B3 Penetrating* | 3 | 12 | *S2 Moderate* | 4 | 44 | | - |
| *+/- Perianal disease* | 5 | | *S3 Severe* | 4 | 44 | | - |
| **Clinical Activity** | | | | | | | |
| | **Median** | **IQR** | | **Median** | **IQR** | | |
| HBI | 6 | 5 | | | | | - |
| SCCAI | | | | 7 | 4.5 | | - |
| Partial Mayo Score | | | | 4 | 4.5 | | - |
| **Previous Treatment Exposure** | | | | | | | |
| | **n** | **%** | | **n** | **%** | | |
| Steroids | 7 | 29 | | 6 | 55 | | 0.26 |
| Azathioprine | 6 | 25 | | 0 | 0 | | 0.15 |
| Infliximab | 3 | 13 | | 3 | 27 | | 0.65 |
| Adalimumab | 4 | 16 | | 0 | 0 | | 0.28 |
| Vedolizumab | 0 | 0 | | 2 | 18 | | 0.09 |
| Ustekinumab | 2 | 8 | | 0 | 0 | | 1 |
| Tofacitinib | 0 | 0 | | 1 | 9 | | 0.31 |
| Filgotinib | 0 | 0 | | 1 | 9 | | 0.31 |
| Upadacitinib | 1 | 4 | | 0 | 0 | | 1 |
| **Biochemical parameters** | | | | | | | |
| | **Median** | **IQR** | | **Median** | **IQR** | | |
| Faecal Calprotectin (μg/g) | 580 | 911 | | 620 | 860 | | 0.31 |
| CRP mg/L | 5 | 9 | | 8.5 | 12 | | 0.32 |
| **Baseline Endoscopic Scores** | | | | | | | |
| | **Median** | **IQR** | | **Median** | **IQR** | | |
| SES-CD | 7.5 | 8.25 | | | | | - |
| UCEIS | | | | 4.5 | 3 | | - |

**Table S4.** Demographics and clinical characteristics of the Scotland – New in-house 16S rRNA sequencing dataset.

### Figures

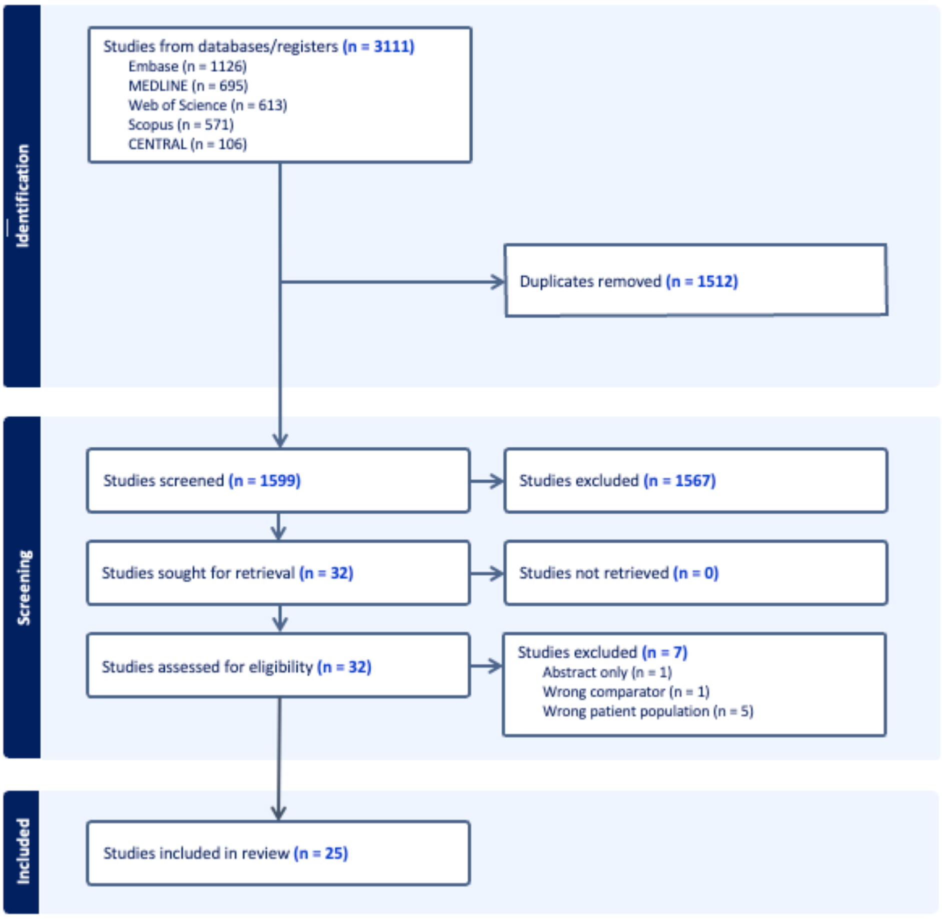

**Figure S1.** PRISMA diagram

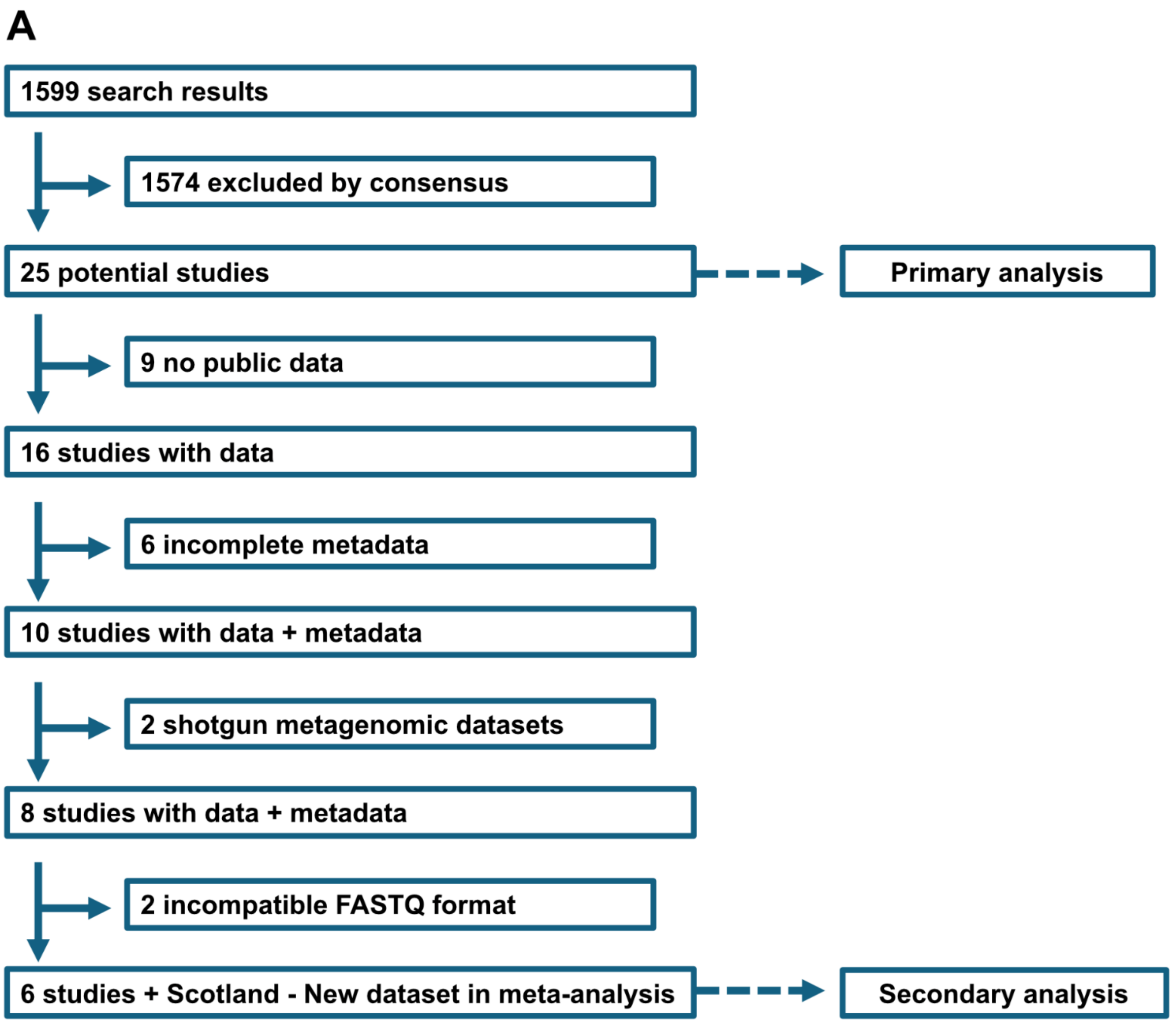

**Figure S2. Data retrieval**

(A) Flowchart of the study selection process. Out of a total 25 potential studies, 6 were included in the final secondary meta-analysis

| Study or Subgroup | Undifferentiated IBD Mean | SD | Total | Control Mean | SD | Total | Weight | Std. mean difference IV, Random, 95% CI |
|---|---|---|---|---|---|---|---|---|
| Abdelbary 2022 | 2.983333333 | 0.395395726 | 14 | 3.23 | 0.259966022 | 14 | 4.5% | -0.72 [-1.48 , 0.05] |
| DeClercq 2025 | 4.346666667 | 0.561070047 | 160 | 4.623333333 | 0.493741642 | 160 | 8.6% | -0.52 [-0.75 , -0.30] |
| Elmaghrawy 2023 | 4.62 | 0.501339925 | 146 | 4.66 | 0.433995756 | 156 | 8.6% | -0.09 [-0.31 , 0.14] |
| Elzayat 2023 | 3.53 | 0.46904724 | 40 | 3.646666667 | 0.299882662 | 40 | 6.9% | -0.29 [-0.73 , 0.15] |
| Hu 2022 | 2.3381 | 0.7061 | 41 | 2.166666667 | 0.516771856 | 21 | 6.2% | 0.26 [-0.27 , 0.79] |
| Kang 2023 | 3.1592 | 0.3853 | 302 | 3.316666667 | 0.330970169 | 100 | 8.6% | -0.42 [-0.65 , -0.19] |
| Molinero 2022 | 4.073333333 | 0.283801143 | 10 | 4.426666667 | 0.254472007 | 11 | 3.5% | -1.26 [-2.22 , -0.31] |
| Park 2022 | 3.4917 | 0.4084 | 38 | 3.493333333 | 0.264319836 | 19 | 6.0% | -0.00 [-0.56 , 0.55] |
| Qi 2021 | 4.2755 | 0.7709 | 22 | 4.273333333 | 0.616520306 | 8 | 4.2% | 0.00 [-0.81 , 0.81] |
| Raisanen 2023 | 2.04 | 0.1775 | 9 | 2.23 | 0.060176394 | 81 | 4.4% | -2.40 [-3.18 , -1.62] |
| Said 2014 | 3.4 | 0.1 | 35 | 3.4 | 0.1 | 24 | 6.2% | 0.00 [-0.52 , 0.52] |
| Somineni 2021 | 2.6068 | 0.3419 | 127 | 2.636666667 | 0.365857503 | 30 | 7.3% | -0.09 [-0.48 , 0.31] |
| Xia 2022 | 2.2954 | 0.5022 | 43 | 2.683333333 | 0.217203597 | 18 | 5.8% | -0.87 [-1.44 , -0.30] |
| Xu 2023 | 4.22 | 0.483417583 | 22 | 4.056666667 | 0.429318158 | 21 | 5.6% | 0.35 [-0.25 , 0.95] |
| Xun 2018 | 5.545 | 0.5695 | 67 | 5.52 | 0.636755668 | 25 | 6.7% | 0.04 [-0.42 , 0.50] |
| Zhang 2020 | 4.7716 | 0.5964 | 60 | 4.753333333 | 0.497399993 | 31 | 7.0% | 0.03 [-0.40 , 0.47] |
| **Total (Wald[a])** | | | **1136** | | | **759** | **100.0%** | **-0.31 [-0.53 , -0.08]** |

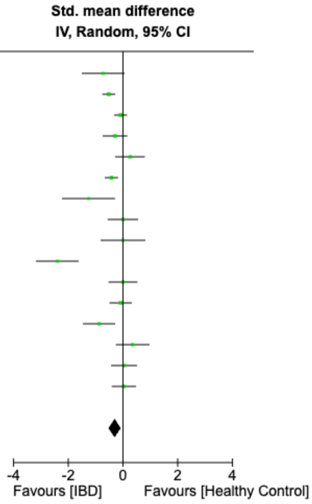

Test for overall effect: Z = 2.69 (P = 0.007)

Heterogeneity: Tau² (DL[b]) = 0.14; Chi² = 62.49, df = 15 (P < 0.00001); I² = 76%

**Footnotes**

[a]CI calculated by Wald-type method.

[b]Tau² calculated by DerSimonian and Laird method.

**Figure S3.** Random effects meta-analysis of the standardised mean difference of salivary Shannon diversity comparing 1136 patients with IBD with 759 controls.

| | | ALPHA DIVERSITY | | | | | | BETA DIVERSITY | | | | |
|---|---|---|---|---|---|---|---|---|---|---|---|---|
| **REF** | Study | Chao1 | Shannon | Simpson | Observed OTU's | Pielou's eveness | Other | Bray-Curtis dissimilarity | Weighted UniFrac | Unweighted UniFrac | Jaccard similarity | Other |
| **51** | Elmaghrawy 2022 | ↔ | ↔ | ↔ | | | | | | | | + ✚ |
| **52** | Abdelbary 2022 | ↔ | ↓ | ↔ | | | * ↔ | ✚ | ✚ | | | |
| **53** | Shin 2022 | | | | | | | | | | | |
| **54** | Somineni 2021 | ↔ | ↔ | ↔ | | | ** ↔ | ↔ | | | | |
| **55** | Elzayat 2023 | ↓ | ↔ | ↔ | ↓ | | * ↓ | ↔ | | | | |
| **56** | Räisänen 2023 | ↔ | ↔ | ↔ | | | | | | | | |
| **57** | Imai 2021 | | | | | | | | | | ✚ | |
| **58** | Kang 2023 | ↓ | ↓ | | | ↔ | | ✚ | | | ✚ | ++ ✚ |
| **59** | Hu 2021 | | ↔ | ↔ | | | | ✚ | | | | |
| **60** | Goel 2023 | ↔ | | ↔ | | | | | | | | ++ ✚ |
| **61** | Sohn 2023 | | | | ↓ | | | | | ↔ | | |
| **62** | Xu 2023 | ↑ | ↔ | ↔ | | | *** ↑ | ✚ | | | | |
| **63** | Zhang 2020 | ↓ | ↔ | ↔ | ↓ | | **** ***** ↓ | | ✚ | ✚ | | |
| **64** | Qi 2021 | ↔ | ↔ | ↔ | ↔ | | * ↔ | | ✚ | | | |
| **65** | Kelsen 2015 | | | | ↓ | | | | ✚ | ✚ | | |
| **66** | Xia 2022 | ↓ | ↓ | | | | | | | ✚ | | |
| **67** | Said 2014 | ↔ | ↔ | ↔ | ↔ | ↔ | * ****** ↔ | | ✚ | ✚ | | |
| **68** | Molinero 2022 | | ↓ | ↔ | ↓ | | | ↔ | | | | |
| **69** | Hu 2022 | | ↔ | | | | | ✚ | | | | |

| | | | | | | | | | | | | |
|---|---|---|---|---|---|---|---|---|---|---|---|---|
| **70** | Monleon-Getino 2023 | ↔ | ↔ | ↔ | ↔ | ↔ | ******* ↔ | ± ✚ | | | | |
| **71** | Xun 2018 | ↔ | ↔ | ↔ | ↔ | | * ***** ↔ | ± ✚ | ± ✚ | ± ✚ | | |
| **72** | Park 2022 | ↔ | ↔ | ↔ | | | * ******* ↔ | ↔ | | | | |
| **73** | DeClercq 2025 | ↓ | ↓ | ↔ | ↓ | | ***** ↓ | ✚ | | ✚ | | + ✚ |
| **74** | Sun 2024 | ↑ | | | | | | | ✚ | | | |
| **75** | Xu 2025 | ↔ | ↔ | | | | | | | | | + ✚ |

| | |
|---|---|
| ↓ Significantly decreased alpha diversity in IBD | ✚ Significant difference in beta diversity reported |
| ↑ Significantly increased alpha diversity in IBD | ↔ No significant difference in beta diversity reported |
| ↔ No Significant difference in alpha diverity | |

Other
*ACE
**Alpha index
***Richness Index
****Good’s coverage
*****Phylogenetic diversity
******Fisher’s alpha index
*******Jackknife2
+ Aitchison’s distance
++ ThetaYC dissimilarity
± Significant difference in CD only, not UC

**Figure S4.** Summary of reported changes in alpha and beta diversity metrics from reported studies.

| Study or Subgroup | Crohn's disease Mean | SD | Total | Control Mean | SD | Total | Weight | Std. mean difference IV, Random, 95% CI |
|---|---|---|---|---|---|---|---|---|
| DeClercq 2025 | 4.353333333 | 0.659596516 | 65 | 4.623333333 | 0.493741642 | 160 | 16.3% | -0.49 [-0.78 , -0.20] |
| Elzayat 2023 | 3.53 | 0.46904724 | 40 | 3.646666667 | 0.299882662 | 40 | 10.7% | -0.29 [-0.73 , 0.15] |
| Hu 2022 | 2.3381 | 0.7061 | 41 | 2.166666667 | 0.516771856 | 21 | 8.5% | 0.26 [-0.27 , 0.79] |
| Kang 2023 | 3.213333333 | 0.299948249 | 127 | 3.316666667 | 0.330970169 | 100 | 17.6% | -0.33 [-0.59 , -0.06] |
| Qi 2021 | 4.38 | 0.905688076 | 12 | 4.273333333 | 0.616520306 | 8 | 3.7% | 0.13 [-0.77 , 1.02] |
| Said 2014 | 3.4 | 0.1 | 21 | 3.4 | 0.1 | 24 | 7.3% | 0.00 [-0.59 , 0.59] |
| Somineni 2021 | 2.59 | 0.384463957 | 87 | 2.636666667 | 0.365857503 | 30 | 11.5% | -0.12 [-0.54 , 0.29] |
| Xia 2022 | 2.2954 | 0.5022 | 43 | 2.683333333 | 0.217203597 | 18 | 7.6% | -0.87 [-1.44 , -0.30] |
| Xun 2018 | 5.496666667 | 0.614611305 | 13 | 5.52 | 0.636755668 | 25 | 6.0% | -0.04 [-0.71 , 0.63] |
| Zhang 2020 | 4.7716 | 0.5964 | 60 | 4.753333333 | 0.497399993 | 31 | 10.9% | 0.03 [-0.40 , 0.47] |
| **Total (Wald[a])** | | | **509** | | | **457** | **100.0%** | **-0.22 [-0.41 , -0.04]** |

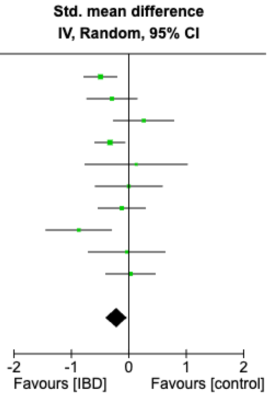

Test for overall effect: Z = 2.34 (P = 0.02)

Heterogeneity: Tau² (DL[b]) = 0.03; Chi² = 14.89, df = 9 (P = 0.09); I² = 40%

**Footnotes**

[a]CI calculated by Wald-type method.

[b]Tau² calculated by DerSimonian and Laird method.

**Figure S5.** Forest plot of salivary Shannon diversity of patients with CD versus control

| Study or Subgroup | Ulcerative Colitis Mean | SD | Total | Control Mean | SD | Total | Weight | Std. mean difference IV, Random, 95% CI |
|---|---|---|---|---|---|---|---|---|
| DeClercq 2025 | 4.36 | 0.515728299 | 64 | 4.623333333 | 0.493741642 | 160 | 18.7% | -0.52 [-0.82 , -0.23] |
| Kang 2023 | 3.12 | 0.433554792 | 175 | 3.316666667 | 0.330970169 | 100 | 19.9% | -0.49 [-0.74 , -0.24] |
| Molinero 2022 | 4.073333333 | 0.283801143 | 10 | 4.426666667 | 0.254472007 | 11 | 6.1% | -1.26 [-2.22 , -0.31] |
| Qi 2021 | 4.15 | 0.593402391 | 10 | 4.273333333 | 0.616520306 | 8 | 6.3% | -0.19 [-1.13 , 0.74] |
| Said 2014 | 3.4 | 0.1 | 14 | 3.4 | 0.1 | 24 | 10.0% | 0.00 [-0.66 , 0.66] |
| Somineni 2021 | 2.643333333 | 0.222989671 | 40 | 2.636666667 | 0.365857503 | 30 | 13.9% | 0.02 [-0.45 , 0.50] |
| Xu 2023 | 4.22 | 0.483417583 | 22 | 4.056666667 | 0.429318158 | 21 | 11.1% | 0.35 [-0.25 , 0.95] |
| Xun 2018 | 5.556666667 | 0.563619206 | 54 | 5.52 | 0.636755668 | 25 | 13.9% | 0.06 [-0.41 , 0.54] |
| **Total (Wald[a])** | | | **389** | | | **379** | **100.0%** | **-0.23 [-0.51 , 0.04]** |

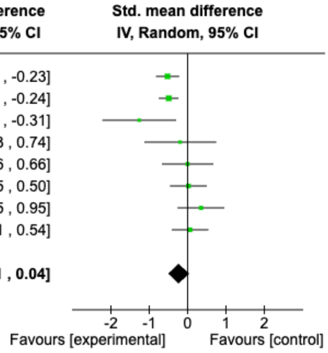

Test for overall effect: Z = 1.68 (P = 0.09)

Heterogeneity: Tau² (DL[b]) = 0.08; Chi² = 17.63, df = 7 (P = 0.01); I² = 60%

**Footnotes**

[a]CI calculated by Wald-type method.

[b]Tau² calculated by DerSimonian and Laird method.

**Figure S6.** Forest plot of salivary Shannon diversity of patients with UC versus control

| Study or Subgroup | IBD - High Activity Mean | SD | Total | Control Mean | SD | Total | Weight | Std. mean difference IV, Random, 95% CI |
|---|---|---|---|---|---|---|---|---|
| Elmaghrawy 2023 | 4.39 | 0.815814716 | 52 | 4.66 | 0.433995756 | 102 | 26.2% | -0.46 [-0.79 , -0.12] |
| Molinero 2022 | 4.073333333 | 0.283801143 | 10 | 4.426666667 | 0.254472007 | 11 | 13.4% | -1.26 [-2.22 , -0.31] |
| Park 2022 | 3.556666667 | 0.4004846 | 19 | 3.493333333 | 0.264319836 | 19 | 19.4% | 0.18 [-0.45 , 0.82] |
| Xia 2022 | 2.256666667 | 0.410889676 | 23 | 2.683333333 | 0.217203597 | 18 | 18.6% | -1.23 [-1.91 , -0.55] |
| Zhang 2020 | 4.67 | 0.725158824 | 29 | 4.753333333 | 0.497399993 | 31 | 22.4% | -0.13 [-0.64 , 0.37] |
| **Total (95% CI)** | | | **133** | | | **181** | **100.0%** | **-0.51 [-0.98 , -0.04]** |

Heterogeneity: Tau² = 0.19; Chi² = 13.15, df = 4 (P = 0.01); I² = 70%
Test for overall effect: Z = 2.14 (P = 0.03)
Test for subgroup differences: Not applicable

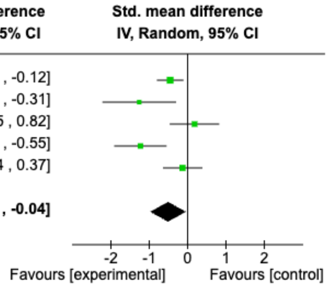

| Study or Subgroup | IBD - Low Activity Mean | SD | Total | Control Mean | SD | Total | Weight | Std. mean difference IV, Random, 95% CI |
|---|---|---|---|---|---|---|---|---|
| Elmaghrawy 2023 | 4.65 | 0.350184937 | 55 | 4.66 | 0.433995756 | 102 | 36.6% | -0.02 [-0.35 , 0.30] |
| Park 2022 | 3.426666667 | 0.416503984 | 19 | 3.493333333 | 0.264319836 | 19 | 19.0% | -0.19 [-0.82 , 0.45] |
| Xia 2022 | 2.34 | 0.598384505 | 23 | 2.683333333 | 0.217203597 | 18 | 19.0% | -0.71 [-1.35 , -0.08] |
| Zhang 2020 | 4.866666667 | 0.435224994 | 31 | 4.753333333 | 0.497399993 | 31 | 25.4% | 0.24 [-0.26 , 0.74] |
| **Total (95% CI)** | | | **128** | | | **170** | **100.0%** | **-0.12 [-0.46 , 0.22]** |

Heterogeneity: Tau² = 0.06; Chi² = 5.58, df = 3 (P = 0.13); I² = 46%
Test for overall effect: Z = 0.68 (P = 0.50)
Test for subgroup differences: Not applicable

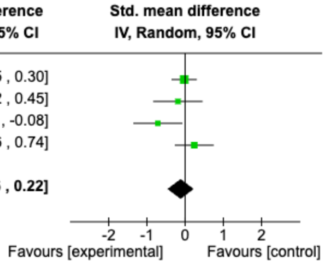

**Figure S7.** Forest plot of salivary Shannon diversity of IBD patients with high disease activity and low disease activity, versus control.

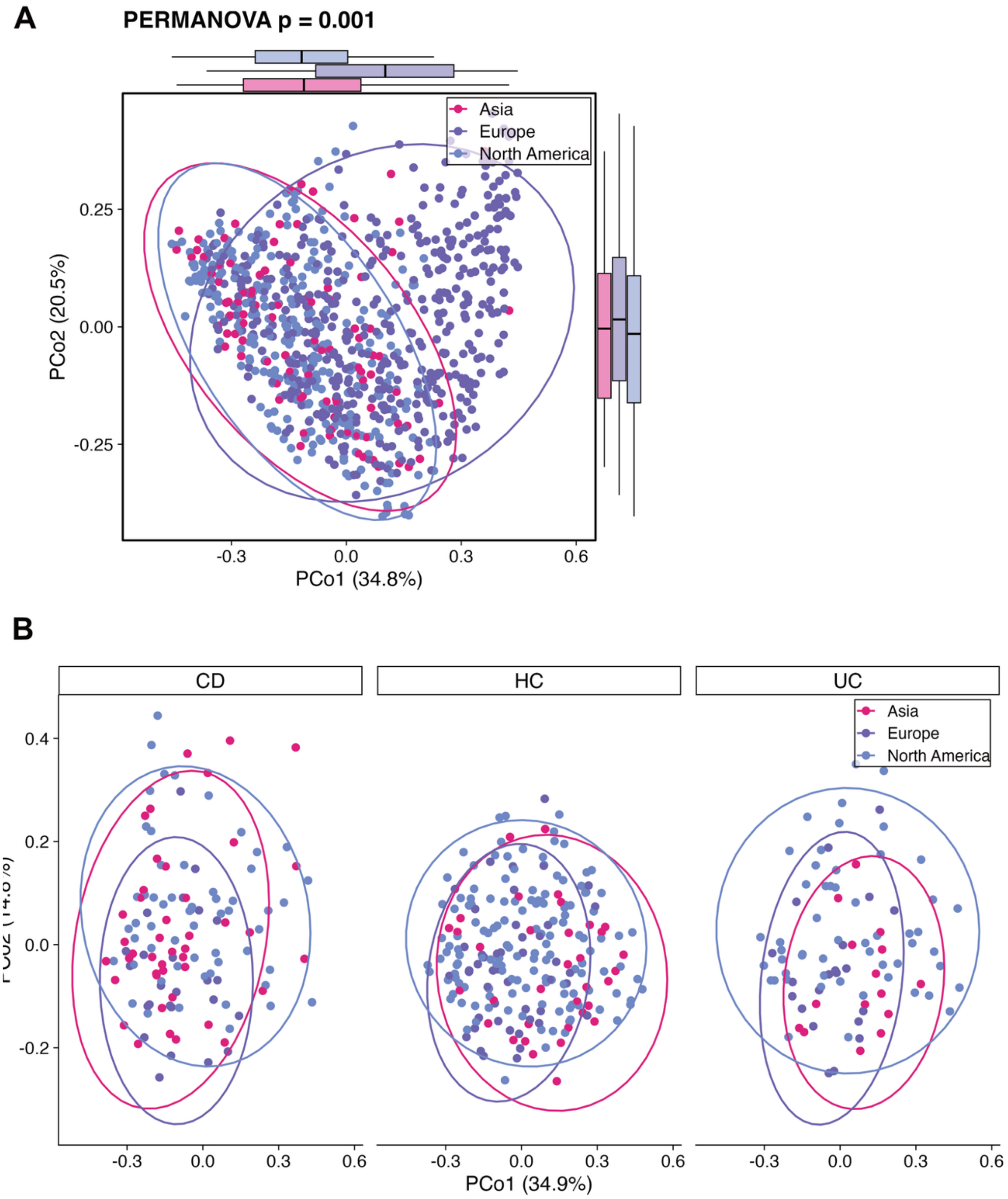

**Figure S8. Oral microbiota beta diversity across geographic locations**

(A–B) Bray-Curtis principal coordinates analysis from geographical groups and PERMANOVA analysis. Colours indicate disease group and ellipses represent 95% confidence bounds around group centroids. (A) represents all samples, and (B) represents IBD subtype stratification.

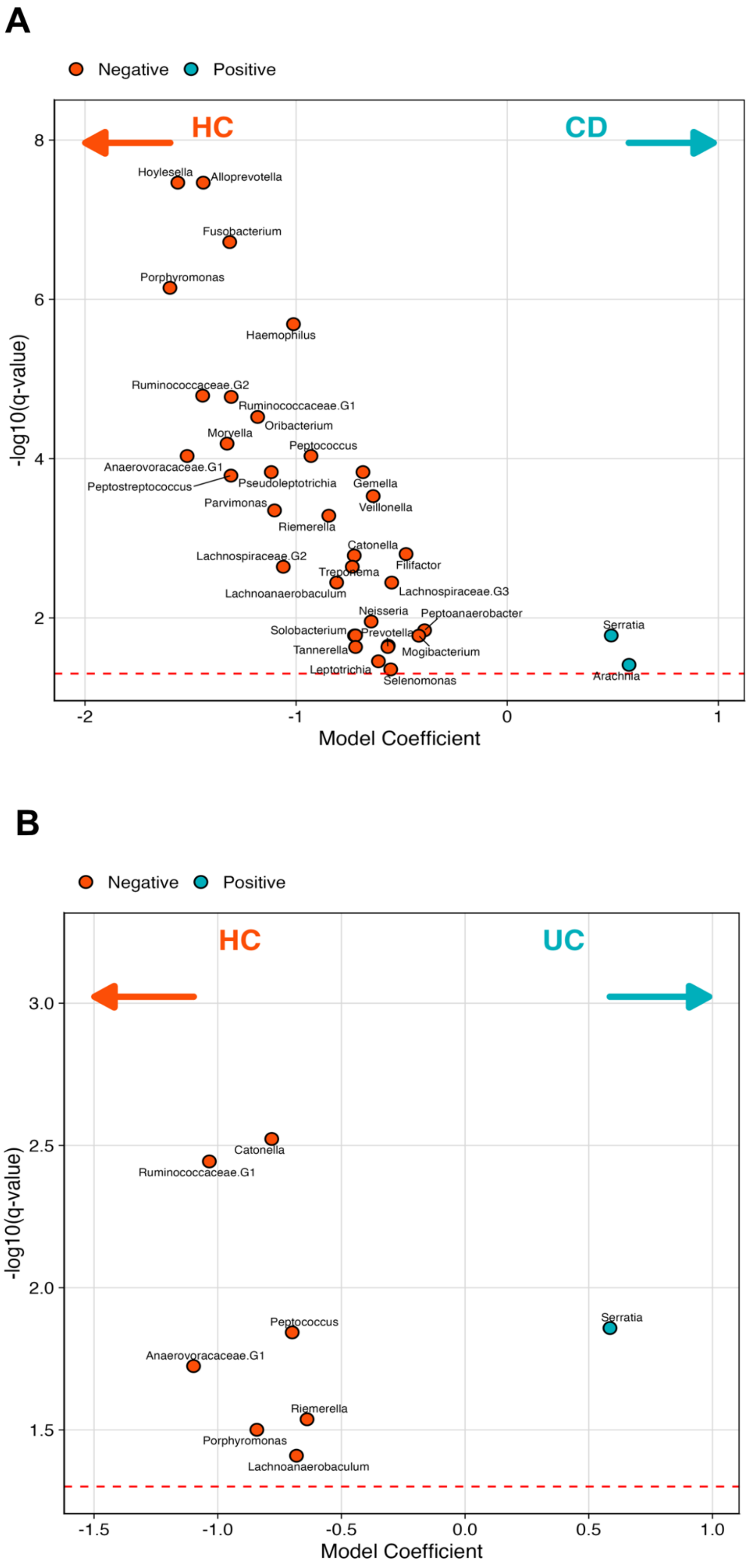
A
Negative
Positive
HC
CD
Hoylesella
Alloprevotella
Fusobacterium
Porphyromonas
Haemophilus
Ruminococcaceae.G2
Ruminococcaceae.G1
Oribacterium
Moryella
Peptococcus
Anaerovoracaceae.G1
Peptostreptococcus
Pseudoleptotrichia
Gemella
Veillonella
Parvimonas
Riemerella
Catonella
Lachnospiraceae.G2
Filifactor
Treponema
Lachnoanaerobaculum
Lachnospiraceae.G3
Neisseria
Peptoanaerobacter
Solobacterium
Prevotella
Tannerella
Mogibacterium
Leptotrichia
Selenomonas
Serratia
Arachnia
-log10(q-value)
Model Coefficient
B
Negative
Positive
HC
UC
Catonella
Ruminococcaceae.G1
Peptococcus
Serratia
Anaerovoracaceae.G1
Riemerella
Porphyromonas
Lachnoanaerobaculum
-log10(q-value)
Model Coefficient

**Figure S9. Altered taxonomic profiles in IBD-subtypes**

Volcano plots displaying MaAsLin2 coefficients (x-axis) of differentially abundant genera in (A) CD and (B) UC using HC as the reference group. Positive coefficients indicate the genus is associated with higher levels in CD/UC (blue), while negative values indicate higher levels in HC (red). $-Log_{10}$-transformed q values are shown on the y axis (Benjamini-Hochberg method). Plot shows only significantly associated taxa ($q < 0.05$).
